# Identifying novel metabolomics risk factors for lacunar stroke and vascular cognitive impairment

**DOI:** 10.1101/2025.03.11.25323678

**Authors:** Wen Hui Ng, Elnara Aghakishiyeva, Hugh S. Markus, Eric L. Harshfield

## Abstract

**Background:** Lacunar stroke results from cerebral small vessel disease (SVD) and is a major cause of vascular dementia and cognitive decline. We evaluated the associations of metabolites with lacunar stroke, neuroimaging markers of SVD, and cognition to help elucidate SVD pathogenesis.

**Methods:** In a clinical cohort of 1,456 MRI-confirmed lacunar stroke cases and 952 controls, we assayed 250 metabolites using nuclear magnetic resonance spectroscopy. We investigated the association of metabolites with lacunar stroke, MRI markers of SVD, and cognitive impairment. We also conducted metabolite genome-wide association analyses and applied Mendelian randomization (MR) to investigate causality.

**Results:** We identified 211 metabolites (84%) that were significantly associated with lacunar stroke. MR analyses identified significance evidence to support causal associations of two of these metabolites, glycine and %C/Total in M-LDL, with two lacunar stroke subtypes, ILI and MLI/LA, respectively. Five metabolites (Total S-HDL, S-HDL concentration, PL in S-HDL, %CE/Total in L-LDL, and lactate) were significantly associated with executive functioning/processing speed, but their causality could not be evaluated. MR analyses also identified evidence to support causal associations of ω-3/Total FA with two diffusion tensor imaging markers of SVD, which were not possible to evaluate in observational analyses. Glycine and %C/Total in M-LDL exhibited the most robust and consistent associations with lacunar stroke across subtypes both the observational and MR analyses.

**Conclusions:** Our findings reveal novel risk factors and emphasize the importance of metabolomics in unravelling metabolic pathways involved in the pathogenesis of lacunar stroke and SVD and cognitive decline.

**Clinical Perspective:** *What is new?:*

- In a clinical cohort of MRI-confirmed lacunar stroke cases and controls, we found robust and consistent evidence across observational and causal inference analyses that glycine and the proportion of cholesterol within medium LDL were significantly associated with two lacunar stroke subtypes, emphasizing the importance of metabolomics in helping to elucidate SVD pathogenesis.

*What are the clinical implications?:*

- These findings highlight the role of metabolites in affecting lacunar stroke and cognitive decline.
- Metabolomics can help guide the development of targeted interventions aimed at preventing stroke and mitigating the progression of cognitive decline.

## Introduction

Cerebral small vessel disease (SVD) is a progressive cerebrovascular disorder affecting the small perforating arteries, arterioles, capillaries, and small veins in the brain.^1^ These small vessels are crucial for maintaining blood flow to the sub-surface brain structure and facilitating optimal functioning of the brain’s most metabolically active white and deep grey matter networks.^1^ SVD is a key mechanism linking stroke and dementia, particularly vascular dementia,^2^ and is responsible for lacunar stroke and most deep intracerebral hemorrhages, resulting in considerable public health burden.^3,4^ The clinical consequences of SVD are diverse, ranging from subtle cognitive impairments and gait disturbances to severe cognitive decline and recurrent strokes.^5^ SVD can be characterized by neuroimaging features shown on MRI, including lacunar infarcts, cerebral microbleeds, white matter hyperintensities (WMH), perivascular spaces, and brain atrophy.^6^

SVD can be divided into amyloid-related and non-amyloid-related sporadic subcortical SVD. The latter subtype, usually related to hypertension, is the topic of this study. Several pathological processes have been described in sporadic SVD, including lipohyalinosis, a diffuse abnormality of the small vessels, and atheroma affecting both the perforating and intracerebral arteries. Early pathological studies delineated two distinct types of sporadic SVD, one with single larger lacunar infarcts due to microatheroma, and one with multiple smaller lacunar infarcts due to lipohyalinosis.^7^ Radiological studies have shown that the latter subtype is often associated with confluent WMH, referred to as leukoaraiosis,^8^ and the two subtypes have distinct cardiovascular risk profiles.^9^

SVD pathogenesis remains poorly understood, partly due to the difficulty in visualizing and investigating such small vessels *in vivo*, and until recently, it received little attention as an important stroke mechanism.^10^ Consequently, no effective treatments or preventive measures for SVD exist. Elucidating the pathophysiology of SVD and uncovering novel risk factors are crucial for guiding development of targeted interventions to prevent stroke and mitigate cognitive decline.

Metabolomics profiling is a promising technique for identifying novel risk factors in SVD to better understand metabolic pathways underlying disease pathogenesis and enhance prediction of disease progression and severity.^11^ This method uses techniques such as nuclear magnetic resonance (NMR) spectroscopy to identify and quantify metabolites with high sensitivity and specificity. The comprehensive metabolite measurements and detailed characterization of metabolic phenotypes allows determination of metabolic arrangements underlying disease pathogenesis and discovery of therapeutic markers and identification of novel biomarkers to monitor and diagnose disease.^12^ Metabolomics has been successfully applied in a number of cardiovascular and neurological diseases,^13,14^ but there have been few studies in SVD. Previous studies have shown associations of metabolites with MRI SVD features in a large community population^15^ and in small cohort studies,^11,16^ but no metabolomics studies exist for large cohorts of lacunar stroke patients and there is limited data on metabolite associations with vascular cognitive impairment (VCI). Metabolites could provide insights into biochemical activities and disease-related pathways within the body, uncovering molecular mechanisms underlying SVD and cognitive decline and potentially serving as useful biomarkers or therapeutic targets.

In this study, we compared metabolite concentrations in both lacunar stroke patients and stroke-free controls in two longitudinal, prospectively recruited, multicenter studies of MRI-confirmed lacunar stroke in the UK (DNA Lacunar 1 and 2). In addition, we analyzed associations of metabolites with lacunar stroke subtypes, imaging markers of SVD, and cognition, allowing the detailed investigation of metabolic pathways underlying lacunar stroke and SVD. We used a triangulation approach to synthesize evidence from observational and causal inference analyses and identify the most promising biomarkers for lacunar stroke.

## Methods

The study is reported following the STROBE (Strengthening the Reporting of Observational Studies in Epidemiology) guidelines for observational and Mendelian randomization studies (**Supplemental Material**).

## Data availability

Recruitment to the DNA Lacunar 2 study is ongoing. Investigators may request access to anonymized individual patient data after recruitment to the study is completed. The authors will share the data with qualified investigators whose proposal for data use has been approved by an independent review committee.

## Study population

DNA Lacunar 1 (DL1) and DNA Lacunar 2 (DL2) are prospective studies of patients with MRI-confirmed lacunar stroke recruited from stroke centers throughout the UK. DL1 recruited 1,030 patients of European ancestry aged ≤70 years with lacunar stroke from 72 specialist stroke centers throughout the UK between 2002 and 2012.^17^ DL2, for which recruitment is currently ongoing, had recruited 1,319 MRI-confirmed lacunar stroke patients as of July 2023 from 54 hospitals across the UK.^18^ Both studies had the same inclusion criteria except that DL1 only recruited patients who had experienced a stroke prior to age 70. Lacunar stroke was identified based on the presence of a clinical lacunar syndrome with an anatomically compatible lesion on MRI, defined as a diffusion-weighted imaging (DWI) positive lesion for acute infarcts, and a cavitated lacune in an anatomically corresponding lesion for cases imaged >3 weeks after the acute event.^17^ The median time from event to MRI was 5 days. Each patient underwent comprehensive stroke investigations including brain MRI, carotid artery imaging and ECG, as well as echocardiography where necessary. Exclusion criteria included stenosis >50% in the extracranial or intracranial cerebral vessels, history of carotid endarterectomy, high or moderate probability of cardioembolic stroke sources defined by the Trial of Org 10172 Acute Stroke Therapy (TOAST) classification criteria,^19^ cortical infarcts on MRI, subcortical infarcts >15mm in diameter, monogenic cause of stroke, or other specific causes of stroke.^17^ All clinical histories and MRI scans were reviewed centrally by an experienced physician to confirm eligibility.

DL1 also recruited 1,973 unrelated hospital controls of European ancestry without cerebrovascular disease from general practices in the same geographical area by random sampling.^17^ Both cases and controls completed standardized clinical assessments and a questionnaire. Brain MRI was not conducted for the controls. Blood was drawn from cases and controls, including serum and plasma, which was centrifuged immediately and stored at –80°C. The consent date was used as an approximation for the date of blood draw for the metabolomics assay since samples were usually taken on the date of consent.

DL1 was approved by the Multi-Centre Research Ethics Committee (04/MRE00/36). DL2 received approval from the East of England–Cambridge Central Research Ethics Committee (16/EE/0201). Informed consent was obtained from all participants in both studies.

### Review of MRI

All MRI scans, performed clinically at each recruiting site, were reviewed centrally to confirm eligibility. As described previously,^20^ modalities included DWI sequences to identify acute infarcts, fluid-attenuated inversion recovery (FLAIR) and/or T1 sequences to count old lacunar infarcts, and FLAIR and/or T2 sequences to identify WMH. Susceptibility-weighted imaging and gradient echo sequences were not mandatory but were used to count cerebral microbleeds if available. A lacune was defined as a round or ovoid subcortical fluid-filled cavity of 3-15 mm in diameter. Microbleeds were counted using the Brain Observer Microbleed Rating Scale (BOMBS), with a microbleed defined as a focal area of very low intensity <10 mm in diameter.^21^ The Fazekas scale was used to grade the extent of WMH on T2-FLAIR images. WMHs were graded by one of three neurologists, who underwent training and passed a standardized assessment prior to reading scans. Cohen’s kappa inter-rater reliability test showed good to very substantial agreement (average kappa=0.788).^20^ The MRI scans were also used to classify lacunar stroke cases into subtypes. Isolated lacunar infarcts (ILI) and multiple lacunar infarcts (MLI) were defined as 1 or >1 lacunar infarct, respectively, without confluent WMH (Fazekas grade <2). Leukoaraiosis (LA) was defined as having confluent WMH (Fazekas grade ≥2) with or without MLI. MLI and LA were combined in accordance with previous studies since they have a shared pathology.^22^

### Cognitive assessment

DL2 lacunar stroke cases were assessed with the Brief Memory and Executive Test (BMET).^20,23^ This is a short cognitive screening tool, designed and validated to be sensitive for the detection of VCI in SVD patients,^23^ which is freely available at www.bmet.info. It includes a number of sub-scores, each ranked on an age normative scale from 0 to 2, which are used to form a total score that ranges from 0 to 16.^23^ A BMET total score of ≤13 has been previously validated to define the presence of VCI.^23^ The test performance can be subdivided into two major cognitive domain clusters, orientation/memory (O/M) and executive functioning/processing speed (EF/PS), each with a score ranging from 0 to 8. The O/M subdomain score was calculated from the tasks for orientation, five-item repetition, five-item recall, and five-item recognition memory, while the EF/PS subdomain score was calculated from the tasks for letter-number matching, motor sequencing, letter sequencing, and number-letter sequencing.^23^

### Metabolomics profiling

Serum samples from 2,500 participants were selected: 673 DL1 cases, 827 DL2 cases, and 1,000 DL1 controls. All selected cases had lacunar stroke confirmed on central review of imaging. Ineligible cases and withdrawn participants were excluded. Serum samples were prioritized for selection amongst participants that had sufficient sample volume (minimum 350 µL). Samples in 0.5 mL tubes in –80 °C freezers were transferred from storage trays and boxes to twenty-seven 96-well microtube racks. Samples were arranged with up to 25 DL1 cases, 31 DL2 cases, and 38 DL1 controls on each rack, with at least 2 empty wells for QC samples.

Metabolic biomarker profiling was performed at Nightingale Health’s laboratories in Finland in September 2023. Serum samples were shipped on dry ice and high-throughput proton nuclear magnetic resonance (NMR) spectroscopy was used to obtain measurements of 250 metabolites for each sample with high precision and accuracy (previously shown to have a median intra-class coefficient of 0.99).^24,25^ All samples were processed on a single spectrometer. Data processing involved advanced proprietary algorithms that translate the NMR spectral data into absolute quantifications of metabolites.^24^ The platform measured a wide range of biomarkers including amino acids, apolipoproteins, cholesterol and cholesteryl esters, fatty acids, phospholipids, triglycerides, and detailed measurements of lipid concentrations and compositions within 14 different lipoprotein subclasses^24^ (**Table S1**).

### Statistical analyses

#### Observational analyses

To ensure approximately normal distributions, prior to statistical analyses a generalized log transformation was applied to each metabolite and the concentrations were rescaled using mean centering and dividing by the standard deviation of each metabolite across participants. Due to high repeatability and negligible batch effects of the biomarker profiling platform and since all samples from DL1 and DL2 were analyzed on a single mass spectrometer, batch correction was not necessary. We excluded samples with insufficient sample volume or measurements below the limit of quantification.

Presence of confluent WMH was defined as Fazekas grade ≥2, and BMET total score ≤13 indicated evidence of VCI. Age-normalized scores for the BMET O/M and EF/PS subdomains were analyzed as continuous outcomes, and the directions were reversed so that a higher score on each subdomain indicated evidence of cognitive impairment.

In our primary analyses, we studied the association of metabolites with lacunar stroke and with the ILI and MLI/LA subtypes by constructing logistic regression models with adjustment for age and sex. Within lacunar stroke cases (i.e. excluding controls), we also analyzed the association of each metabolite with presence of WMH, CMBs, and VCI using logistic regression, and with the BMET O/M and EF/PS subdomain scores using linear regression, all adjusted for age and sex.

As sensitivity analyses, we conducted regression analyses for the association of each metabolite with the above outcomes with further adjustment for hypertension, smoking status, alcohol consumption, body mass index, hyperlipidemia, and type 2 diabetes. We also conducted a sensitivity analysis with further adjustment for both vascular risk factors and medications prescribed for stroke treatment, including antihypertensives, diabetic drugs, lipid-lowering drugs, antiplatelets, and anticoagulants. Furthermore, we conducted a sensitivity analysis within lacunar stroke cases adjusted for the number of days between the stroke event and the date of blood sample collection.

We also performed a principal component analysis (PCA) of metabolite levels and assessed the association of the top principal components, which explained the greatest proportion of the variance in metabolite concentrations, with each of the above outcomes with adjustment for age and sex.

Observational analyses were conducted in R v4.4.1. A false discovery rate (FDR) threshold of 5% was used to account for testing associations with 250 metabolites for each outcome.

#### Causal inference analyses

To complement the observational analyses, we conducted a genome-wide association analysis for each of the 250 metabolites using genetic data from DL1 and DL2, and then we performed two-sample Mendelian randomization analyses (MR) using the independent SNPs for each metabolite to examine whether there was evidence to support a causal relationship of metabolites with lacunar stroke and MRI markers of SVD. Further details of genotyping, imputation, quality control, and genome-wide association analyses are provided in the **Supplemental Methods**.

Genetic association summary statistics for white matter hyperintensities (WMH) (n=55,291), lower fractional anisotropy (FA) (n=36,533), mean diffusivity (MD) (n=36,460), peak width of skeletonized mean diffusivity (PSMD) (n=36,012), cerebral microbleeds (CMB) (n=3,556 cases; n=22,306 controls), MRI-confirmed lacunar stroke (n=3,199), ILI (n=1,133), and MLI/LA (n=1,658) were obtained from published studies, which used data from UK Biobank, the Cohorts of Heart and Aging Research in Genomic Epidemiology (CHARGE) Consortium, and DNA Lacunar 1 and 2.^26,27^ All summary statistics were for individuals of European ancestry except for CMB, which included 3% of participants from other ancestries.

Palindromic genetic variants with ambiguous allele frequencies and genetic variants with potential strand issues that could not be resolved were excluded. All independent variants associated with the metabolites were harmonized with the outcome data to ensure that the effect estimates of each variant with both the exposure and the outcome corresponded to the same effect allele. Proxy variants identified from the 1000 Genomes EUR reference population were used for a small number of exposure–outcome pairs where the lead SNPs after clumping were not found in the outcome dataset. Primary analyses were conducted using the inverse-variance weighted (IVW) method, or the Wald ratio method when only one genetic variant was available to be used as an instrumental variable.

Sensitivity analyses were performed including the weighted median, simple and weighted mode, and MR-Egger regression methods, which require different assumptions to be satisfied thus supporting the credibility of inferred causal claims.^28^ Cochran’s Q statistic obtained from the IVW method was used to diagnose heterogeneity and outliers. MR-Egger intercepts were used to evaluate whether the results were influenced by directional pleiotropy.

For the metabolites with evidence to support significant causal associations, we performed reverse MR analyses to examine whether there was evidence to suggest that the outcomes may have a causal effect on the metabolite levels.

MR analyses were conducted in R using the TwoSampleMR v0.6.8 package with an FDR threshold of 5% to control for multiple testing.

#### Interaction analyses

As an additional sensitivity analysis, we examined whether there was significant evidence of interaction with vascular risk factors, namely hypertension, smoking status, alcohol consumption status, body mass index, hyperlipidemia, Type 2 diabetes status, and age category. We examined whether there was significant interaction for metabolites that were significantly associated with lacunar stroke or its subtypes in both the observational and MR analyses, and for metabolites that were significantly associated with the BMET EF/PS subdomain in the observational analyses.

## Results

### Participant characteristics

Metabolomics measurements that passed quality control were returned for 2,408 participants— 645 DL1 cases, 811 DL2 cases, and 952 DL1 controls. Cases from DL1 and DL2 were combined. A flow chart of the sample selection process is shown in **Figure 1**, and details of participant characteristics are summarized in **Tables 1**, **2**, and **3**. A comparison of the characteristics for selected participants with DL1 and DL2 participants who were not included in the analysis is provided in **Table S2**, with no significant differences between groups for nearly all characteristics indicating minimal likelihood of selection bias.

**Figure 1.**
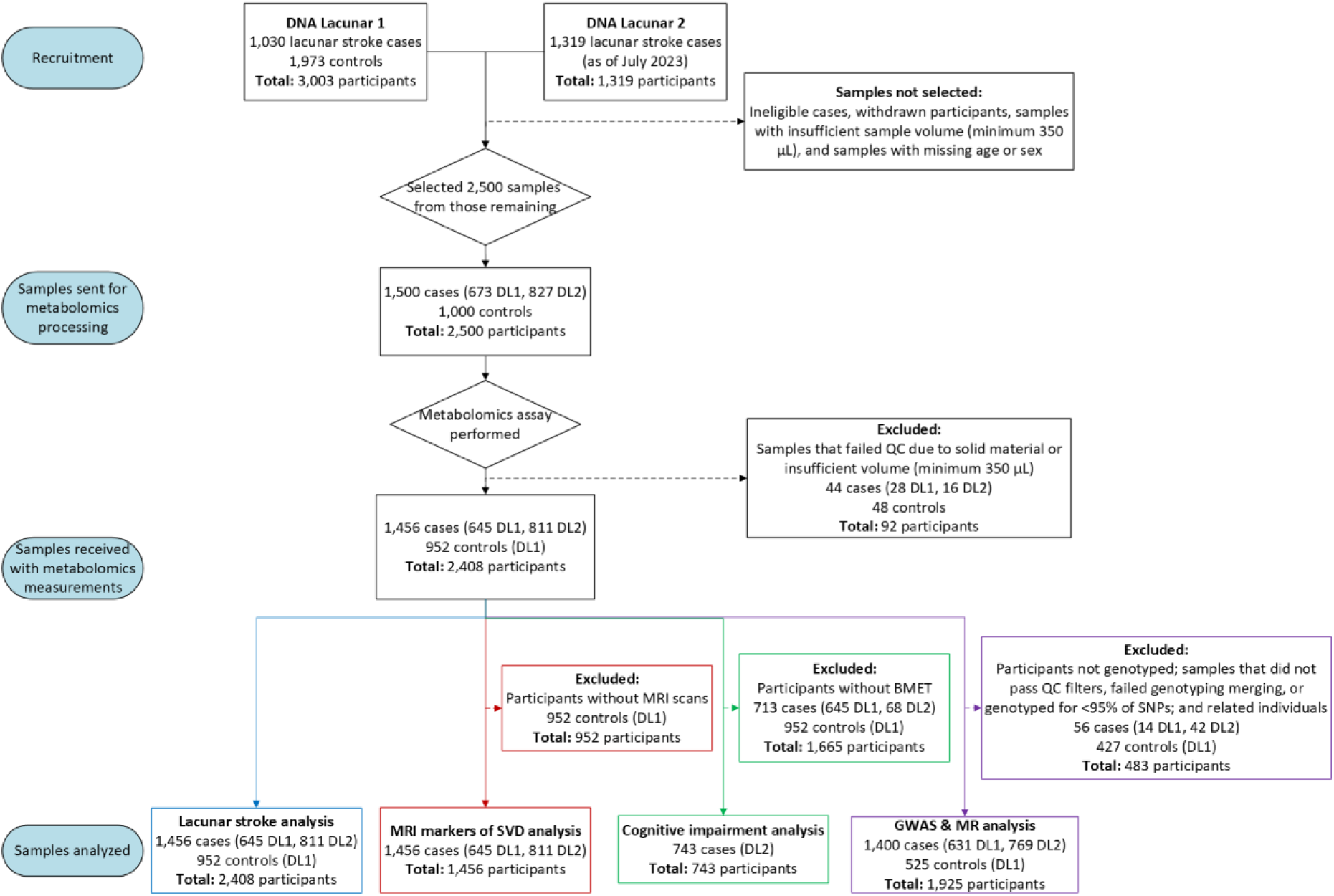
Flow diagram of sample selection process. Abbreviations: BMET: Brief Memory and Executive Test; EF/PS: executive functioning/processing speed; ILI: isolated lacunar infarcts; LA: leukoaraiosis; MLI: multiple lacunar infarcts; O/M: orientation/memory; MR: Mendelian randomization; SNP: single nucleotide polymorphism; SVD: small vessel disease; QC: quality control.

**Table 1.** Demographics, vascular risk factors, and medications in lacunar stroke cases and controls.

| Characteristic | Total n | Cases (n=1,456) | Controls (n=952) | P-value |
| --- | --- | --- | --- | --- |
| Age at recruitment (years) | 2,408 | 61.92 (11.54) | 57.49 (6.25) | <0.001 |
| Male sex | 2,408 | 962 (66.1%) | 595 (62.5%) | 0.0803 |
| Hypertension | 2,408 | 1,046 (71.8%) | 443 (46.5%) | <0.001 |
| Hyperlipidemia | 2,395 | 955 (65.7%) | 479 (50.9%) | <0.001 |
| Type 2 diabetes | 2,408 | 283 (19.4%) | 62 (6.5%) | <0.001 |
| Current smoker | 2,407 | 419 (28.8%) | 131 (13.8%) | <0.001 |
| Alcohol consumption | 2,395 | 966 (66.5%) | 749 (79.5%) | <0.001 |
| Body mass index (kg/m<sup>2</sup>) | 2,384 | 28.45 (6.09) | 27.38 (5.26) | <0.001 |
| Antihypertensives | 2,408 | 621 (42.7%) | 196 (20.6%) | <0.001 |
| Diabetic drugs | 2,408 | 234 (16.1%) | 43 (4.5%) | <0.001 |
| Lipid-lowering drugs | 2,395 | 415 (28.5%) | 159 (16.9%) | <0.001 |
| Antiplatelets | 2,395 | 914 (62.8%) | 106 (11.3%) | <0.001 |
| Anticoagulants | 2,398 | 45 (3.1%) | 14 (1.5%) | 0.0245 |

Participant characteristics are shown for lacunar stroke cases and controls from DNA Lacunar 1 and 2 with metabolomics data. Mean and SD are shown for continuous variables, while frequency and percentage are shown for categorical variables. *P*-values are from *t*-tests for continuous variables and from chi-squared tests for categorical variables, showing a statistical comparison of cases vs controls. Abbreviations: SD: standard deviation.

**Table 2.** Demographics, vascular risk factors, and medications in lacunar stroke subtypes (ILI and MLI/LA) and comparison with controls.

| Characteristic | Total n (ILI / MLI/LA) | ILI (n=610) | P-value | MLI/LA (n=842) | P-value |
| --- | --- | --- | --- | --- | --- |
| Age at recruitment (years) | 610 / 842 | 58.35 (10.86) | 0.0747 | 64.50 (11.35) | <0.001 |
| Male sex | 610 / 842 | 397 (65.1%) | 0.3269 | 563 (66.9%) | 0.0602 |
| Hypertension | 610 / 842 | 386 (63.3%) | <0.001 | 657 (78.0%) | <0.001 |
| Hyperlipidemia | 610 / 840 | 391 (64.1%) | <0.001 | 561 (66.8%) | <0.001 |
| Type 2 diabetes | 610 / 842 | 96 (15.7%) | <0.001 | 186 (22.1%) | <0.001 |
| Current smoker | 610 / 841 | 179 (29.3%) | <0.001 | 238 (28.3%) | <0.001 |
| Alcohol consumption | 609 / 840 | 438 (71.9%) | 0.0007 | 525 (62.5%) | <0.001 |
| Body mass index (kg/m<sup>2</sup>) | 600 / 832 | 28.83 (6.28) | <0.001 | 28.18 (5.95) | 0.0028 |
| Antihypertensives | 610 / 842 | 201 (33.0%) | <0.001 | 419 (49.8%) | <0.001 |
| Diabetic drugs | 610 / 842 | 77 (12.6%) | <0.001 | 156 (18.5%) | <0.001 |
| Lipid-lowering drugs | 610 / 840 | 137 (22.5%) | 0.0079 | 277 (33.0%) | <0.001 |
| Antiplatelets | 610 / 842 | 340 (55.7%) | <0.001 | 573 (68.1%) | <0.001 |
| Anticoagulants | 610 / 842 | 11 (1.8%) | 0.7808 | 33 (3.9%) | 0.0023 |

Participant characteristics are shown for lacunar stroke cases according to subtypes (ILI and MLI/LA) from DNA Lacunar 1 and 2 with metabolomics data. Mean and SD are shown for continuous variables, while frequency and percentage are shown for categorical variables. *P*-values are from *t*-tests for continuous variables and from chi-squared tests for categorical variables, showing a statistical comparison of ILI vs controls and MLI/LA vs controls. Abbreviations: ILI: isolated lacunar infarcts; LA: leukoaraiosis; MLI: multiple lacunar infarcts; SD: standard deviation.

Lacunar stroke cases were slightly older than controls (**Table 1**), with a mean age of 61.9 (SD 11.5) for cases and 57.5 (SD 6.3) for controls. 66.1% of cases and 62.5% of controls were male. Cases were significantly more likely to have hypertension (71.8% vs 46.5%), hyperlipidemia (65.7% vs 50.9%), and type 2 diabetes (19.4% vs 6.5%) than controls, and cases were more likely to smoke (28.8% vs 13.8%) but less likely to consume alcohol (66.5% vs 79.5%). Cases also had a slightly higher average body mass index: 28.5 kg/m^2^ (SD 6.1) for cases and 27.4 kg/m^2^ (SD 5.3) for controls.

Within lacunar stroke subtypes, similar participant characteristics were observed for cases with ILI and cases with MLI/LA when compared to controls (**Table 2**). However, cases with MLI/LA were slightly older than cases with ILI, with a mean age of 64.5 (SD 11.4) for MLI/LA and 58.4 (10.9) for ILI. There was also a higher proportion of hypertension (78.0% vs 63.3%) and type 2 diabetes (22.1% vs 15.7%) and a lower proportion of alcohol consumption (62.5% vs 71.9%) in cases with MLI/LA compared to those with ILI.

Cerebral microbleeds were much more common in cases with MLI/LA than in cases with ILI (41.7% vs 9.7%) (**Table 3**). Cases with MLI/LA had worse cognition on average, with a BMET total score of 12.6 (SD 3.7) for cases with MLI/LA and 14.0 (SD 2.6) for cases with ILI. Cases with MLI/LA also had worse cognitive scores on the O/M and EF/PS subdomains than cases with ILI.

**Table 3.** Neuroimaging markers and cognition in lacunar stroke cases and subtypes (ILI and MLI/LA)

| Characteristic | Total n (Cases /<br>ILI / MLI/LA) | Cases (n=1,456) | ILI (n=610) | MLI/LA<br>(n=842) |
| --- | --- | --- | --- | --- |
| WMH (Fazekas score $\geq 2$) | 1,455 / 610 / 842 | 591 (40.6%) | 4 (0.7%) | 587 (69.7%) |
| Presence of CMBs | 718 / 267 / 451 | 214 (29.8%) | 26 (9.7%) | 188 (41.7%) |
| BMET total score (VCI) | 743 / 284 / 459 | 13.13 (3.35) | 13.95 (2.59) | 12.62 (3.65) |
| BMET O/M subdomain | 746 / 283 / 463 | 6.55 (1.88) | 6.92 (1.58) | 6.32 (2.02) |
| BMET EF/PS subdomain | 735 / 283 / 452 | 6.67 (1.98) | 7.06 (1.62) | 6.43 (2.14) |

Participant characteristics are shown for lacunar stroke cases overall and according to subtypes (ILI and MLI/LA) from DNA Lacunar 1 and 2 with metabolomics data. Mean and SD are shown for continuous variables, while frequency and percentage are shown for categorical variables. BMET total score ranges from 0-16 where a score $\leq 13$ is suggestive of VCI. The BMET O/M and EF/PS subdomains each range from 0-8 where a lower score indicates worse cognition. The direction was reversed in regression analyses so that a higher value on the subdomains corresponds to worse cognition. Abbreviations: BMET: Brief Memory and Executive Test; CMB: cerebral microbleeds; EF/PS: executive functioning/processing speed; ILI: isolated lacunar infarcts; LA: leukoaraiosis; MLI: multiple lacunar infarcts; O/M: orientation/memory; SD: standard deviation; VCI: vascular cognitive impairment; WMH: white matter hyperintensities.

### Observational associations with lacunar stroke

An overview of the key findings from the observational and causal inference analyses is shown in **Figure 2**. In the observational regression analyses, we found that 211 metabolites out of 250 (84%) were significantly associated with lacunar stroke at an FDR-corrected threshold of *P* < 0.05 (**Figures 2**, **3** and **4**, **Figure S1**, and **Table S3**). Lower levels of amino acids, ketone bodies, apolipoproteins, fatty acids, cholesterol and cholesteryl esters, multiple lipoprotein subclasses, and multiple relative lipoprotein lipid concentrations, and higher levels of acetoacetate, creatinine, fluid balance, glucose, glycoprotein acetyls (GlycA), 3-Hydroxybutyrate, and other relative lipoprotein lipid concentrations were associated with increased risk of lacunar stroke. Similar associations were also observed within subsets of lacunar stroke cases with ILI and with MLI/LA compared to controls (**Figures S2** and **S3** and **Table S3**).

**Figure 2.**
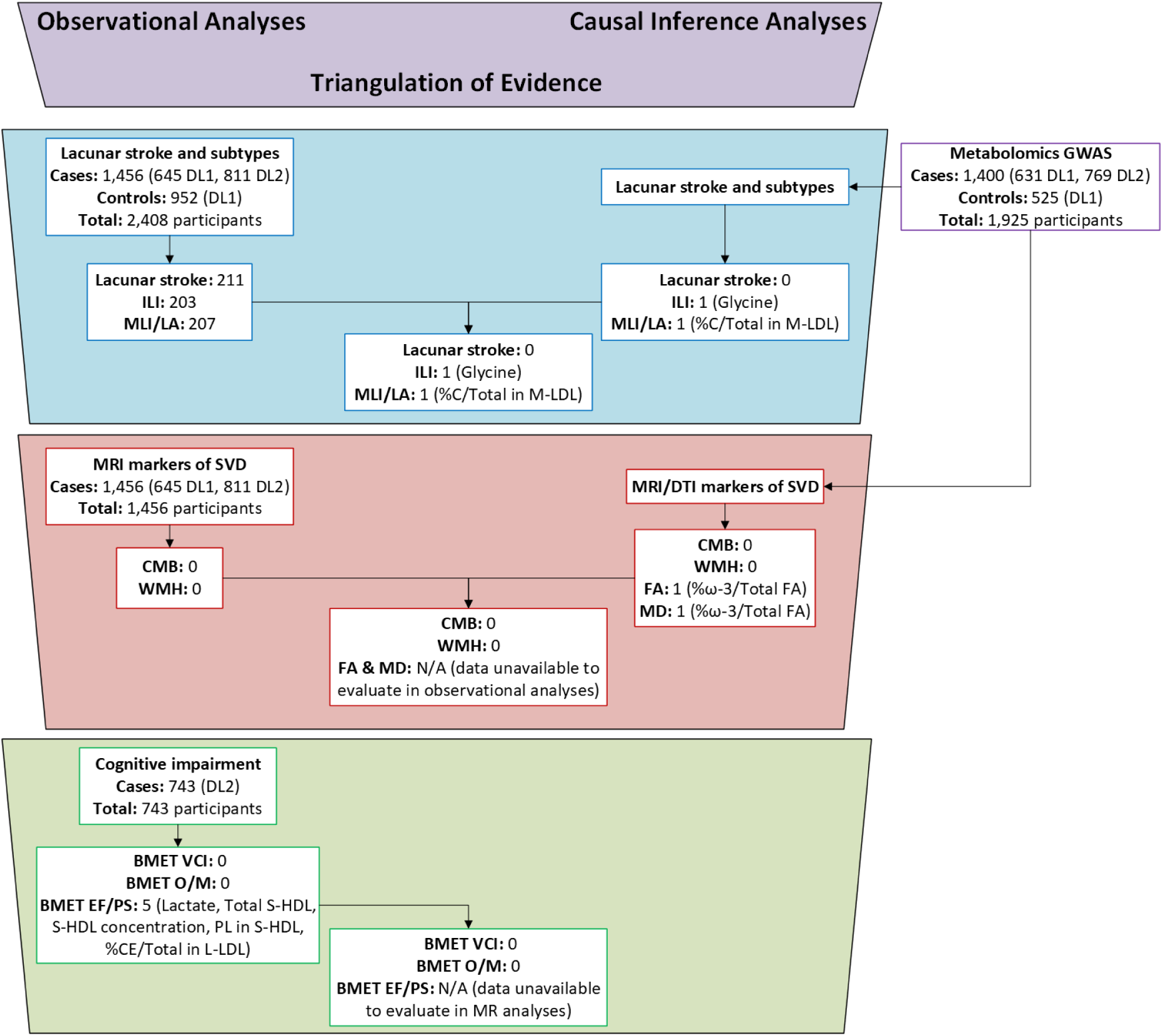
Overview of results. Abbreviations: BMET: Brief Memory and Executive Test; CMB: cerebral microbleeds; DTI: diffusion tensor imaging; EF/PS: executive functioning/processing speed; FA: fractional anisotropy; ILI: isolated lacunar infarcts; LA: leukoaraiosis; MD: mean diffusivity; MLI: multiple lacunar infarcts; MR: Mendelian randomization; O/M: orientation/memory; SNP: single nucleotide polymorphism; SVD: small vessel disease; VCI: vascular cognitive impairment; WMH: white matter hyperintensities.

**Figure 3.**
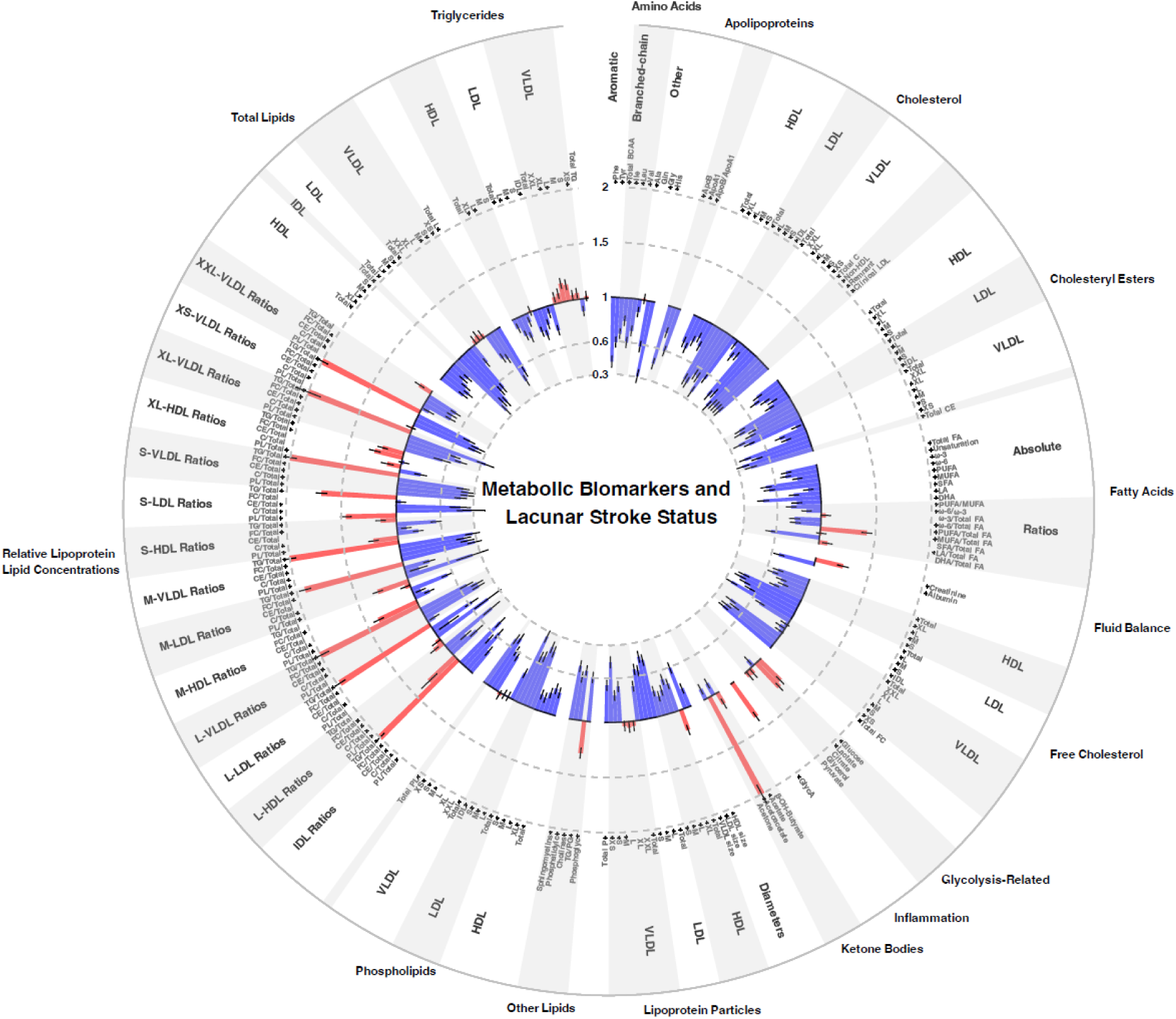
Circular bar plot showing the results of cross-sectional univariate analyses for the association between 250 metabolic biomarkers and lacunar stroke. Abbreviations: FDR: false discovery rate; SD: standard deviation. Odds ratios with 95% confidence intervals for the association with lacunar stroke versus controls are presented per 1-SD increase in metabolite concentration with adjustment for age, sex, hypertension, and recruitment center. Associations shown in red indicate increased odds of lacunar stroke, and in blue indicate reduced odds of lacunar stroke. Associations significant at an FDR-corrected *P* < 0.05 are indicated with an asterisk (*) alongside the names of the respective metabolites.

**Figure 4.**
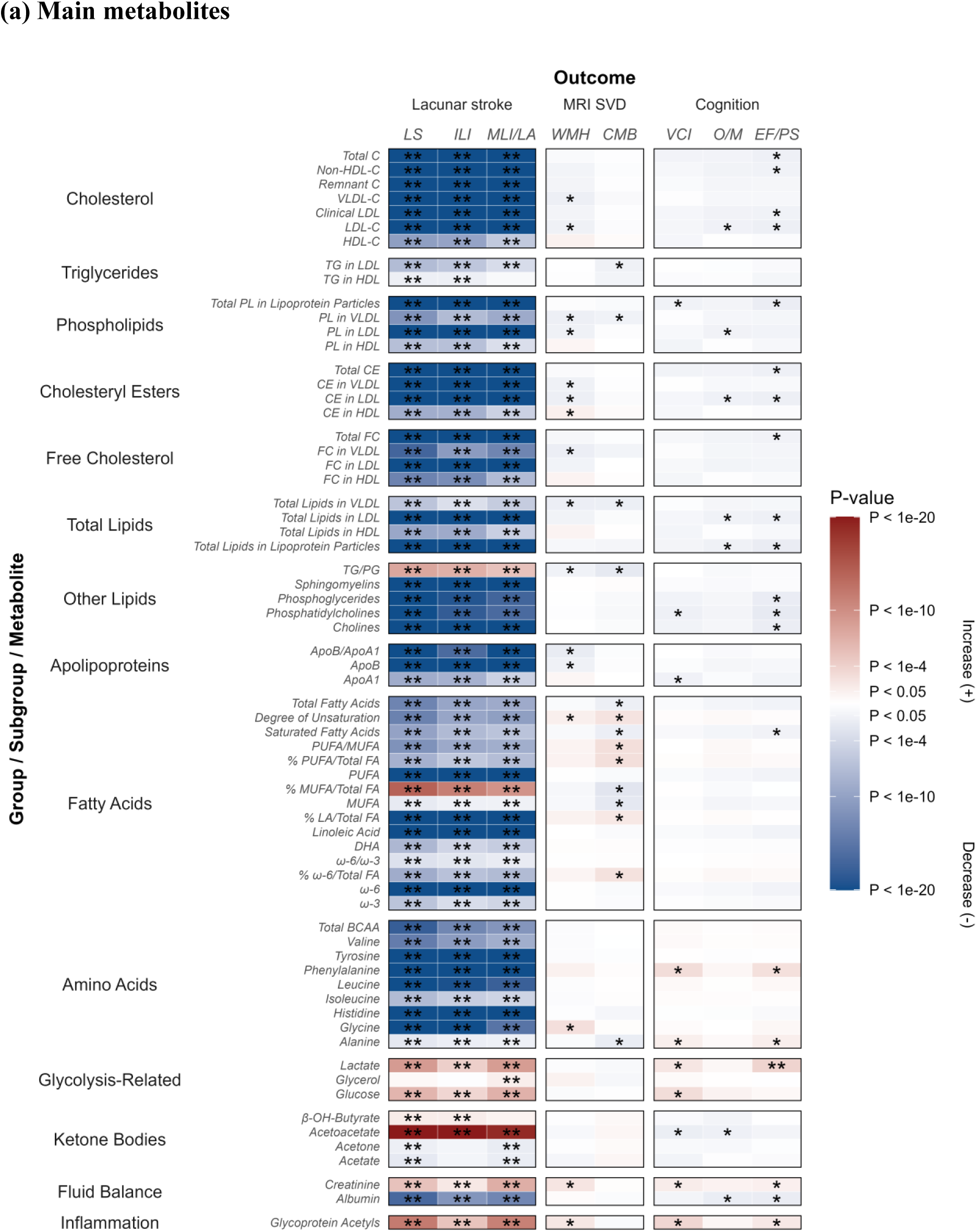
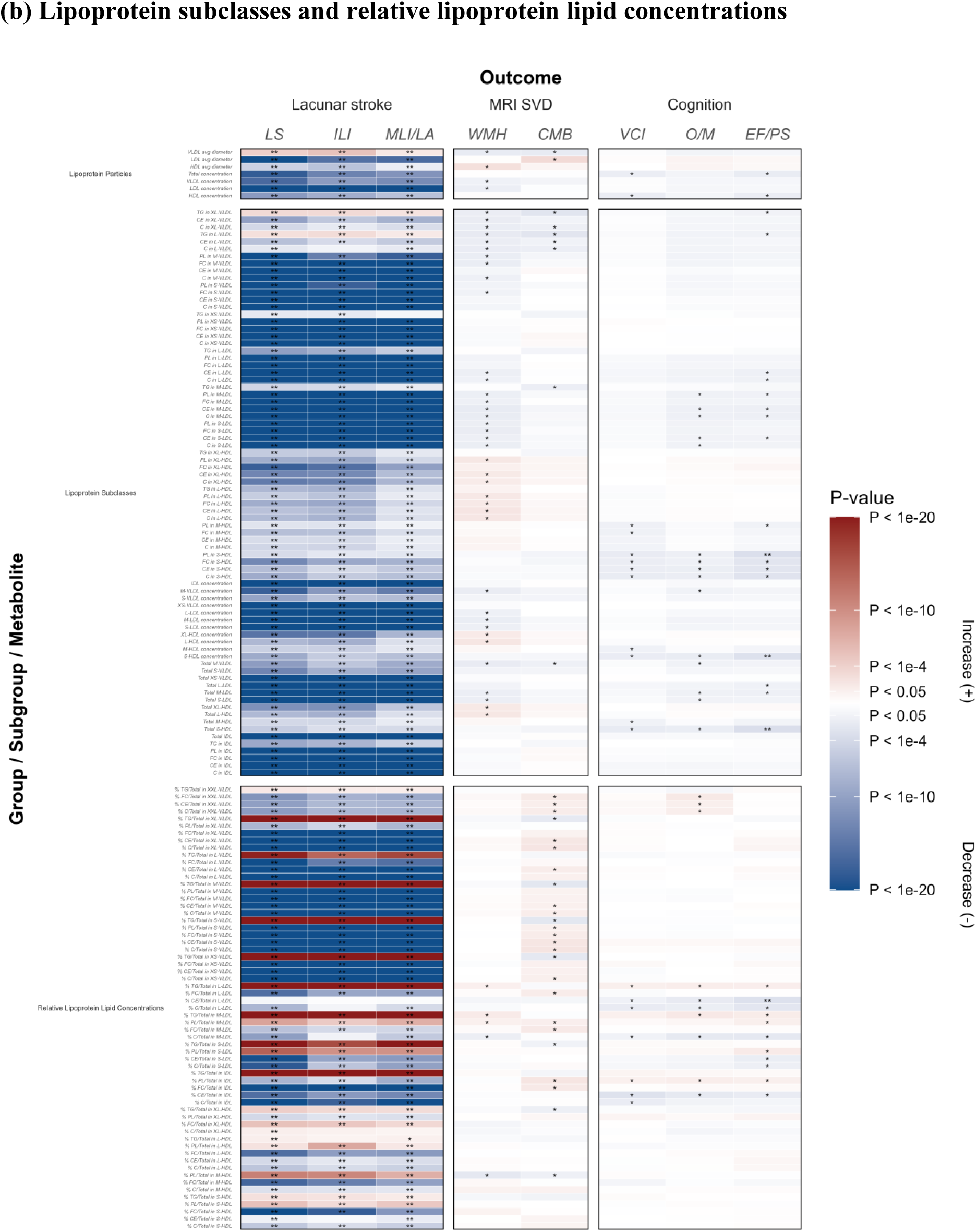
Heat map showing the results of cross-sectional univariate analyses for the association between 250 metabolic biomarkers and lacunar stroke, MRI markers, and cognitive impairment. (a) Main metabolites. (b) Lipoprotein subclasses and relative lipoprotein lipid concentrations. Abbreviations: BMET: Brief Memory and Executive Test; EF/PS: executive functioning/processing speed; FDR: false discovery rate; ILI: isolated lacunar infarcts; LA: leukoaraiosis; MLI: multiple lacunar infarcts; O/M: orientation/memory; SD: standard deviation; SVD: small vessel disease; WMH: white matter hyperintensities. Beta estimates and *P*-values were obtained from logistic or linear regression models adjusted for age and sex. Colors show magnitude and direction of *P*-value for association of metabolite with each outcome. Associations with beta coefficients >1 are represented in red, and associations with beta coefficients <1 are shown in blue. Associations significant at an unadjusted *P* < 0.05 are indicated with a single asterisk (*) and associations significant at an FDR-corrected *P* < 0.05 are indicated with two asterisks (**).

In sensitivity analyses adjusted for additional vascular risk factors and potential confounders, there were still 211 metabolites significantly associated with lacunar stroke after FDR correction. There was a high degree of overlap with the results from the primary analysis, but 28 metabolites, mostly from lipoprotein subclasses, were only associated with lacunar stroke in the fully adjusted model, and 28 metabolites, mostly from lipoprotein subclasses and relative lipoprotein lipid concentrations, were only associated with lacunar stroke in the basic adjustment model **(Figure 2** and **Table S4**). In an additional sensitivity analysis further adjusted for stroke medications, including antihypertensives, diabetic drugs, lipid-lowering drugs, antiplatelets, and anticoagulants, 203 metabolites were significantly associated with lacunar stroke after FDR correction, similarly with a high degree of overlap with results from the primary analysis (**Table S5**).

We also conducted a principal component analysis on the concentrations of 250 metabolites. A scree plot (**Figure S4a**) showed that the first four principal components explained more than 75% of the variance in the metabolite concentrations and the first 22 principal components explained more than 95% of the variance. A scatter plot of the matrix loadings (**Figure S4b**) showed that there was variability in metabolite levels by the overall metabolite classification according to the first and second principal components. Fourteen of the first 22 principal components had significant associations with lacunar stroke after correction for multiple testing (**Figure 2**, **Figure S5,** and **Table S6**). Similar associations were also observed for cases with ILI and with MLI/LA compared to controls.

### Observational associations with neuroimaging markers and cognitive impairment

Within the 1,456 lacunar stroke cases, we conducted cross-sectional univariate analyses of metabolites with two MRI markers of SVD, namely the presence of confluent WMH and cerebral microbleeds, with adjustment for age and sex. Although 75 metabolites and 73 metabolites had suggestive associations (uncorrected *P*<0.05) with WMH and cerebral microbleeds, respectively, none of the associations remained statistically significant after correction for multiple testing (**Figures 2** and **4** and **Table S7**).

Likewise, there were no significant associations with MRI markers in sensitivity analyses adjusted for additional vascular risk factors and potential confounders (**Table S8**), in analyses adjusted for stroke medications (**Table S9**), and analyses adjusted for the duration between the stroke event and the date that the blood sample was taken (**Table S10**).

We also analyzed associations of metabolites with cognitive impairment. Within the 743 lacunar stroke cases from DL2 whom had completed the BMET, the association of 250 baseline metabolic measures with VCI and with scores on the O/M and EF/PS subdomains was assessed in cross-sectional univariate analyses adjusted for age and sex. There were no metabolites that had statistically significant associations with total BMET score after correction for multiple testing (**Figures 2** and **4** and **Table S11**). However, in our separate analyses of the BMET subdomains, we found that five metabolites pertaining to lipoprotein concentrations, lipoprotein ratios, and glycolysis (Total S-HDL, S-HDL concentration, PL in S-HDL, %CE/Total in L-LDL, and lactate) were significantly associated with the EF/PS subdomain.

These five metabolites remained significantly associated with the EF/PS subdomain in sensitivity analyses, including after adjustment for additional vascular risk factors and potential confounders (**Table S12**), adjustment for stroke medications (**Table S13**), and adjustment for the duration between the stroke event and the date of blood sample collection (**Table S14**).

### Causal inference analyses for lacunar stroke and neuroimaging markers

DNA samples were genotyped for 1,958 participants out of the 2,408 individuals with metabolomics data. After excluding samples that did not pass quality control, genetic data were available for 1,925 participants: 631 DL1 cases, 769 DL2 cases, and 525 DL1 controls (**Table S15**).

There were 72 metabolites with at least one genome-wide significant association (**Table S16**). The most pleiotropic genetic loci were *LINC02755*, which was associated with a broad range of lipid traits including triglycerides and fatty acids, and *APOE*, which was associated with multiple fatty acids and lipoproteins.

To complement the observational analyses, we conducted MR analyses for the 72 metabolites with at least one genome-wide significant association. We identified two metabolites with significant evidence of a causal effect with lacunar stroke subtypes at an FDR-corrected threshold of *P* < 0.05, and one metabolite with significance evidence of a causal effect with two diffusion tensor imaging (DTI) markers of SVD (**Figures 2** and **5** and **Table S17**).

**Figure 5.**
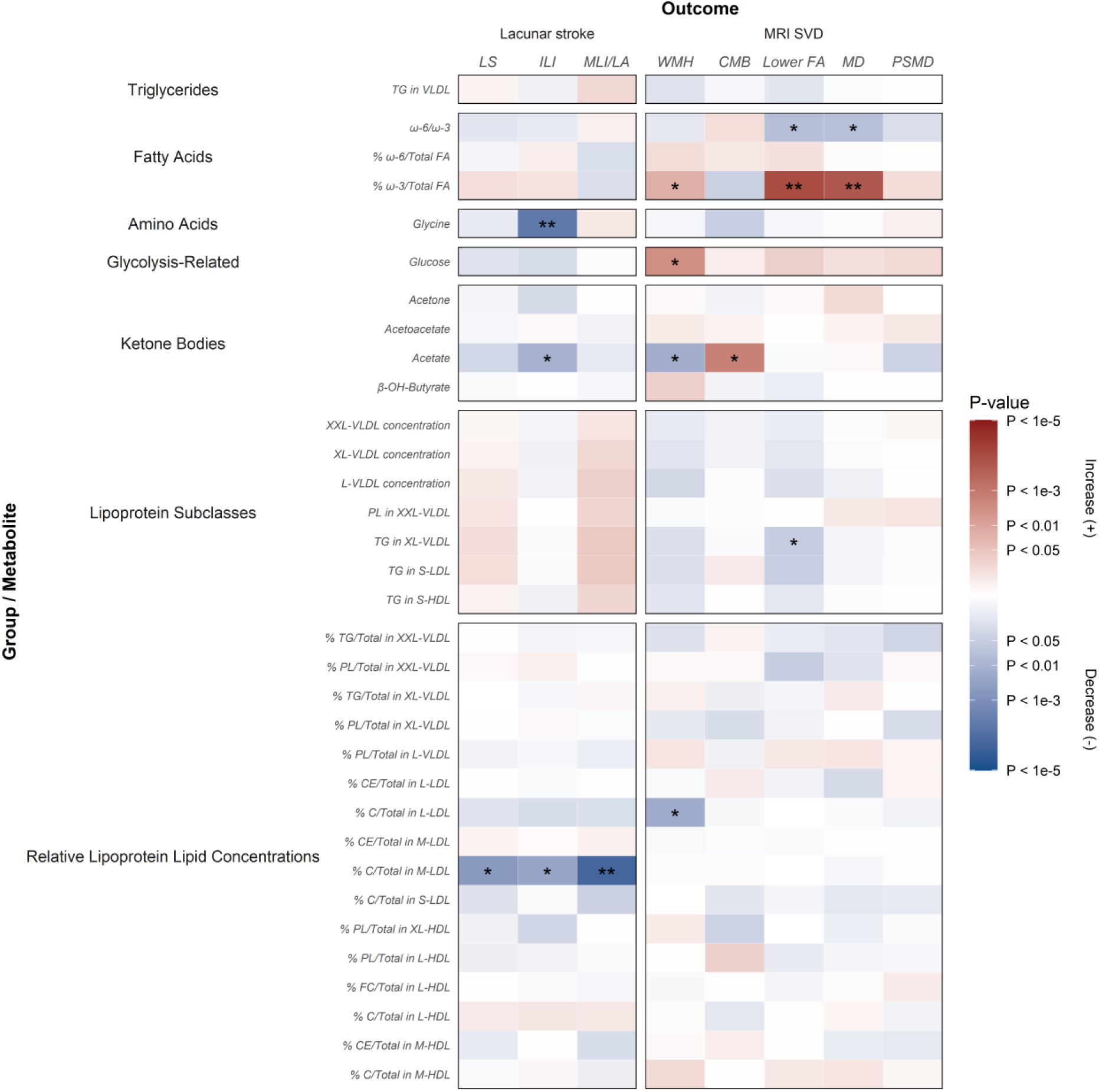
Heat map summarizing the causal estimates from Mendelian randomization analyses for the association of metabolic biomarkers with lacunar stroke and MRI markers of SVD. Abbreviations: CMB: cerebral microbleeds; FA: fractional anisotropy; FDR: false discovery rate; ILI: isolated lacunar infarcts; IVW: inverse-variance weighted; LA: leukoaraiosis; MD: mean diffusivity; MLI: multiple lacunar infarcts; PSMD: peak width of skeletonized mean diffusivity; SD: standard deviation; SVD: small vessel disease; WMH: white matter hyperintensities. Metabolites associated with at least one outcome at *P* < 0.5 using the IVW method are shown. Beta estimates and *P*-values were obtained from two-sample Mendelian randomization analyses using the IVW method, or using the Wald ratio method if only a single SNP was available to be used as an instrumental variable. Colors show magnitude and direction of *P*-value for association of metabolite with each outcome. Associations with beta coefficients >1 are represented in red, and associations with beta coefficients <1 are shown in blue. Associations significant at an unadjusted *P* < 0.05 are indicated with a single asterisk (*) and associations significant at an FDR-corrected *P* < 0.05 are indicated with two asterisks (**).

Genetically determined levels of glycine were significantly associated with a reduced risk of ILI, and genetic predisposition to a higher proportion of cholesterol to total lipids within medium LDL (%C/Total in M-LDL) was significantly associated with a reduced risk of MLI/LA. There was also suggestive evidence (uncorrected *P*<0.05) that genetically determined levels of acetate may reduce the risk of ILI and that genetic predisposition to higher %C/Total in M-LDL may reduce the risk of overall lacunar stroke and ILI, but these associations were not significant after correction for multiple testing.

Genetic predisposition to a higher percentage of omega-3 within total fatty acids (%ω-3/Total FA) was significantly associated with worse white matter damage on DTI, indicated by lowered FA and elevated MD. There was also suggestive evidence that genetically elevated %ω-3/Total FA may also increase WMH volume, but this association was not significant after correction for multiple testing. We also found several other suggestive associations of metabolites with neuroimaging markers of SVD, namely for genetic predisposition to a higher ratio of omega-6 to omega-3 fatty acids (ω-6/ω-3) with lowered FA and with elevated MD, for genetically raised glucose levels with increased WMH volume, for genetically elevated acetate with decreased WMH volume and increased cerebral microbleeds, for genetically determined levels of triglycerides in very large VLDL (TG in XL-VLDL) with elevated FA, and genetic predisposition to a higher proportion of cholesterol to total lipids within large LDL (%C/Total in L-LDL) with decreased WMH volume.

The MR results were directionally concordant across sensitivity analyses (weighted median, simple and weighted mode, MR-Egger) with no evidence of directional or horizontal pleiotropy. We also conducted reverse MR analyses, using lacunar stroke and neuroimaging markers as the exposure and metabolite levels as the outcome, which indicated that the significant associations that we identified were all bidirectional (**Figure 6** and **Table S18**).

**Figure 6.**
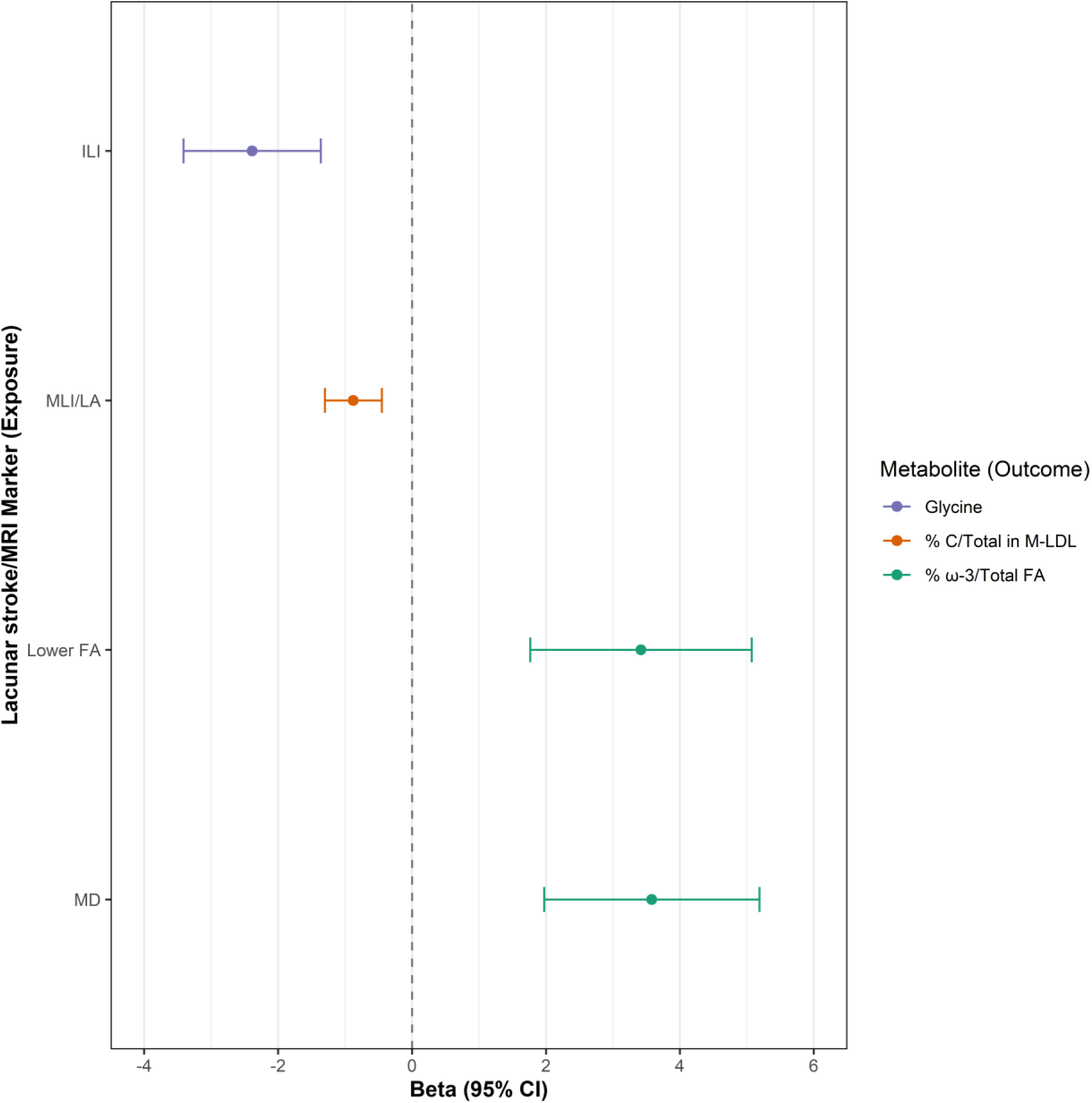
Forest plot summarizing the significant estimates from reverse Mendelian randomization analyses for the association of lacunar stroke and MRI markers of SVD with metabolic biomarkers. Abbreviations: FA: fractional anisotropy; FDR: false discovery rate; ILI: isolated lacunar infarcts; LA: leukoaraiosis; MD: mean diffusivity; MLI: multiple lacunar infarcts; SVD: small vessel disease. Beta estimates and 95% confidence intervals were obtained from two-sample Mendelian randomization analyses using the inverse-variance weighted method.

### Interaction analyses

We identified two metabolites (glycine and %C/Total in M-LDL) that were significantly associated with lacunar stroke subtypes in both the observational and causal inference analyses. To triangulate and validate these findings, we examined whether there was evidence of significant interaction in these associations with vascular risk factors (**Table S19**). We found significant evidence that hypertension, smoking status, body mass index, hyperlipidemia, diabetes status, and age category interacted in the association of %C/Total in M-LDL with MLI/LA. However, there was no evidence that vascular risk factors interacted in the association of glycine with ILI. Likewise, there was no evidence of significant interaction with vascular risk factors for the five metabolites that were significantly associated with the BMET EF/PS subdomain (Total S-HDL, S-HDL concentration, PL in S-HDL, %CE/Total in L-LDL, and lactate).

## Discussion

In this metabolomics study of 2,408 lacunar stroke cases and controls, we conducted observational and causal inference analyses and triangulated the evidence to identify metabolites with concordant associations with lacunar stroke subtypes and neuroimaging markers of SVD. Observational analyses identified 211 metabolites that were significantly associated with lacunar stroke, and MR analyses confirmed evidence of a causal association of genetically elevated glycine with reduced risk of ILI and of genetic predisposition to higher %C/Total in M-LDL with reduced risk of MLI/LA. There were no statistically significant associations of metabolites with neuroimaging markers of SVD in observational analyses, but there was significant evidence of a causal effect of genetically elevated ω-3/Total FA on two DTI markers indicative of white matter damage. We also identified five metabolites pertaining to lipoprotein concentrations, lipoprotein ratios, and glycolysis (Total S-HDL, S-HDL concentration, PL in S-HDL, %CE/Total in L-LDL, and lactate) were significantly associated with the BMET EF/PS subdomain in lacunar stroke cases, which is the subdomain that is characteristically most impaired in SVD;^29^ however, we did not have available genetic data to evaluate whether these associations were likely to be causal. Overall, glycine and %C/Total in M-LDL exhibited the most robust and consistent associations with lacunar stroke across both the observational and MR analyses. These findings highlight possible shared underlying metabolic pathways and illustrate the complex interplay between metabolites in affecting lacunar stroke and cognitive decline.

Glycine is a simple amino acid that has been shown to suppress inflammation and reduce the risk of cardiovascular disease and dementia.^38,39^ Glycine had robust evidence for an inverse association with ILI in both the observational and MR analyses, which has not been previously reported. There was no evidence of interaction with vascular risk factors; however, the reverse MR analyses provided evidence that ILI also had a significant causal effect on glycine concentration, so the associations are bidirectional and it is difficult to disentangle the true nature of the causal relationship. There may be a feedback loop in which metabolite perturbations influence disease susceptibility, and the disease pathogenesis affects metabolite levels.

Acetate showed significant inverse associations with lacunar stroke and MLI/LA in the observational analyses, but in the MR analyses there was only suggestive evidence that the inverse association with ILI may be causal. There was also suggestive evidence in the MR analyses that genetically elevated acetate may lead to decreased WMH volume and increased cerebral microbleeds. Acetate and other short-chain fatty acids have been shown to improve blood-brain barrier integrity and suppress neuroinflammation,^40^ which matches the protective associations that we identified for lacunar stroke and WMH. It is also well-supported that higher levels of glucose lead to larger WMH load through glycation-mediated endothelial damage. Our findings align with existing literature and suggest that glycemic control could be used to reduce WMH volume.

We found strong evidence for a protective effect of the percentage of cholesterol to total lipids in medium LDL (%C/Total in M-LDL) with MLI/LA in both the observational and MR analyses. A previous MR analysis found no association for the absolute quantity of cholesterol in medium LDL (C in M-LDL) with small vessel stroke,^35^ which our study confirmed. However, since we found a robust association for %C/Total in M-LDL, it appears that the ratio of the lipoprotein subfractions within medium LDL-C, rather than the absolute quantity, may have greater impact on lacunar stroke onset. Another MR study found a suggestive but not statistically significant association of %C/Total in M-LDL with small vessel stroke,^36^ but our study may have been able to detect a stronger, statistically significant association since we used MRI-confirmed lacunar stroke, which has improved specificity of stroke subtyping.^18^ A further MR study, which did use MRI-confirmed lacunar stroke as the outcome, found associations of several metabolites with lacunar stroke,^37^ but none that overlapped with the metabolites reported in our study.

The reverse MR analyses showed that the causal relationship between %C/Total in M-LDL and lacunar stroke was also bidirectional, and our interaction analyses suggested the presence of vertical pleiotropy. Several vascular risk factors (hypertension, smoking status, body mass index, hyperlipidemia, diabetes status, and age category) interacted in the association of %C/Total in M-LDL with lacunar stroke and MLI/LA. A related metabolite, %C/Total in L-LDL, had significant inverse associations with lacunar stroke and MLI/LA in the observational analyses, but in the MR analyses there was only suggestive evidence of a causal association with WMH.

Sensitivity analyses indicated that the bidirectional effects that we identified in the reverse MR analyses are not artefacts of pleiotropic pathways, affecting both metabolite levels and stroke risk independently. The MR-Egger intercept tests for horizontal pleiotropy were not statistically significant (*P*>0.05) in most of the forward and reverse MR analyses, and heterogeneity (Cochran’s Q) was generally low, supporting the validity of the causal estimates.

Although we did not have genetic data to evaluate whether the observed associations of the metabolites with the BMET EF/PS subdomain in lacunar stroke cases were likely to be causal, a previous study of UK Biobank participants identified consistent associations of several of the same metabolites, showing that lower levels of Total S-HDL, S-HDL concentration, and %CE/Total in L-LDL were associated with increased white matter structural damage on DTI and increased risk of all stroke and all-cause dementia.^15^ Another study also showed that S-HDL concentration was associated with all-cause dementia and vascular dementia, and that %CE/Total in L-LDL was associated with all-cause dementia and Alzheimer’s disease.^30^ The present study adds to this knowledge by showing that these metabolites are also associated with worse executive function and processing speed in individuals who have experienced a lacunar stroke.

Studies have also developed metabolite scores, such as the MetaboHealth score and a metabolic aging score, which have used a combination of metabolites, including lactate and Total S-HDL, for prediction of all-cause and cause-specific mortality, neurocognitive function, functional independence, and biological aging.^31–33^ Our study showed that lactate is also associated with lacunar stroke, ILI, and MLI/LA, and with worse executive function and processing speed, although there was no evidence to support causality. Lactate plays a significant role in many physiological and pathological processes, including regulation of energy metabolism, immune response, and memory formation, and is thought to potentially be a viable target for treatment of cardiovascular and inflammatory diseases.^34^

Overall, despite many promising findings across a comprehensive set of analyses, glycine and %C/Total in M-LDL were the only metabolites with robust, significant associations in the same direction of effect for lacunar stroke subtypes in both the observational and MR analyses. These two metabolites appear to be credible candidate biomarkers for lacunar stroke, and they each target different subtypes of lacunar stroke.

This study has several important strengths. The analyses were conducted in a unique population, a clinical cohort of MRI-confirmed lacunar stroke cases and controls, to examine the role that metabolites play in lacunar stroke onset and disease progression. Numerous metabolites were identified that are associated with lacunar stroke, highlighting the importance of metabolic pathways in the disease pathophysiology, and indicating areas for further research in risk prediction and screening for potential therapeutic targets.

This study also has several limitations. First, neuroimaging markers were only available in DL2 cases and cognitive scores were only available in a subset of DL2 cases; due to the smaller sample size, these analyses had less power to detect statistically significant associations. Second, cognition was assessed using the BMET, which has been previously validated as being highly sensitive to detect VCI in patients with SVD,^23^ but the test has primarily been designed as a screening tool for VCI and has not been evaluated for comprehensive neuropsychological testing and assessment. Third, an FDR threshold was applied to correct for multiple testing and reduce the likelihood of identifying false-positive associations, but this may have inadvertently discounted some biologically and clinically meaningful associations that did not reach the threshold for statistical significance. Fourth, the observational analyses were cross-sectional since we did not have follow-up data available for clinical endpoints, so longitudinal analyses could not be conducted to evaluate the effect of changes in metabolite levels on long-term risk of cerebrovascular and neurological conditions to better understand disease progression over time. Repeat measures of metabolites would also enable characterization of longitudinal dynamics and how metabolite levels change over the preclinical, acute, and recovery phases of SVD. Fifth, only a small fraction of the 250 metabolites had associations with genetic variants that passed the genome-wide significance threshold in the GWAS, possibly due to the small sample size, greatly limiting the number of genetic instruments available for the MR analyses. Finally, our analyses were conducted in a specific clinical cohort of MRI-confirmed lacunar stroke cases and controls from the UK, so further studies would be required to validate these findings in other patient populations and ancestries.

In this study, we identified 211 metabolites that were significantly associated with lacunar stroke, most of which were also associated with lacunar stroke subtypes, and we identified five metabolites pertaining to lipoprotein concentrations, lipoprotein ratios, and glycolysis that were associated with lower execution function and processing speed. MR analyses identified that glycine and %C/Total in M-LDL had evidence to support potentially causal associations with lacunar stroke and MRI markers of SVD, with concordant associations in the observational and MR analyses. Although further research is needed to validate the findings and discern the biochemical pathways through which metabolites impact lacunar stroke outcomes, this study provides valuable evidence which has helped uncover novel metabolic markers of lacunar stroke and SVD. Ultimately, this could support the development of a metabolite fingerprint that evaluates the risk of SVD and cognitive decline through clinical blood tests.

## Non-standard Abbreviations and Acronyms

BMET: Brief Memory and Executive Test
CMB: Cerebral microbleeds
CT: Computed tomography
DL1: DNA Lacunar 1
DL2: DNA Lacunar 2
DWI: Diffusion-weighted imaging
EF/PS: Executive functioning/processing speed
FA: Fractional anisotropy
FDR: False discovery rate
FLAIR: Fluid-attenuated inversion recovery
GWAS: Genome-wide association study
ILI: Isolated lacunar infarcts
IVW: Inverse-variance weighted
LA: Leukoaraiosis
MD: Mean diffusivity
MLI: Multiple lacunar infarcts
MRI: Magnetic resonance imaging
MR: Mendelian randomization
MS: Mass spectrometry
NMR: Nuclear magnetic resonance
O/M: Orientation/memory
PSMD: Peak width of skeletonized mean diffusivity
QC: Quality control
SVD: Small vessel disease
VCI: Vascular cognitive impairment
WMH: White matter hyperintensities

## Acknowledgements

Members of the DNA Lacunar 1 and 2 Coordinating Centers, Recruitment Centers, and Collaborators are listed in the **Supplemental Material in Appendix 1**.

For the purpose of open access, the authors have applied a Creative Commons Attribution (CC BY) license to any Author Accepted Manuscript version arising from this submission.

## Supplemental Material

Appendix 1

Figures S1-S6

Tables S1-S19

