## Supplemental Material for "Identifying novel metabolomics risk factors for lacunar stroke and vascular cognitive impairment"

#### Table of Contents


##### Appendices

**Appendix 1. Members of DNA Lacunar 1 and 2 Coordinating Centres and Recruiting Centres.**

##### Supplemental Methods

**Detailed Methods. Further details of genome-wide association analyses.**

##### Supplemental Figures

**Supplemental Figure 1. Volcano plot showing the results of cross-sectional univariate analyses for the association between 250 metabolic biomarkers and lacunar stroke.**

**Supplemental Figure 2. Circular bar plot showing the results of cross-sectional univariate analyses for the association of 250 metabolic biomarkers with ILI cases versus controls.**

**Supplemental Figure 3. Circular bar plot showing the results of cross-sectional univariate analyses for the association of 250 metabolic biomarkers with MLI/LA cases versus controls.**

**Supplemental Figure 4. Scree plot and loadings plot from principal component analysis of metabolite levels.**

**Supplemental Figure 5. Forest plot showing the results of cross-sectional univariate analyses for the association of the first twenty-two principal components of metabolite levels with lacunar stroke status, ILI vs controls, and MLI/LA vs controls.**

**Supplemental Figure 6. Scatter plots of genetic associations of metabolic levels with lacunar stroke and neuroimaging markers of SVD for Mendelian randomization association analyses with significant or suggestive associations.**

#### Supplemental Tables

**Supplemental Table 1. Metabolic measures analysed using nuclear magnetic resonance spectroscopy in DNA Lacunar 1 and 2.**

**Supplemental Table 2. Comparison of characteristics of participants from DNA Lacunar 1 and 2 included in analysis and excluded from analysis.**

**Supplemental Table 3. All results for association of lacunar stroke per 1-SD higher baseline metabolite levels with adjustment for age and sex.**

**Supplemental Table 4. All results for association of lacunar stroke per 1-SD higher baseline metabolite levels with adjustment for age, sex, and additional vascular risk factors and potential confounders.**

**Supplemental Table 5. All results for association of lacunar stroke per 1-SD higher baseline metabolite levels with adjustment for age, sex, additional vascular risk factors and potential confounders, and stroke medications.**

**Supplemental Table 6. All results for association of lacunar stroke per 1-SD higher principal component of metabolite levels with adjustment for age and sex.**

**Supplemental Table 7. All results for association of neuroimaging markers of small vessel disease per 1-SD higher baseline metabolite levels with adjustment for age and sex.**

**Supplemental Table 8. All results for association of neuroimaging markers of small vessel disease per 1-SD higher baseline metabolite levels with adjustment for age, sex, and additional vascular risk factors and potential confounders.**

**Supplemental Table 9. All results for association of neuroimaging markers of small vessel disease per 1-SD higher baseline metabolite levels with adjustment for age, sex, additional vascular risk factors and potential confounders, and stroke medications.**

**Supplemental Table 10. All results for association of neuroimaging markers of small vessel disease per 1-SD higher baseline metabolite levels with adjustment for age, sex, and duration between stroke event and date blood sample was taken.**

**Supplemental Table 11. All results for association of cognitive decline per 1-SD higher baseline metabolite levels with adjustment for age and sex.**

**Supplemental Table 12. All results for association of cognitive decline per 1-SD higher baseline metabolite levels with adjustment for age, sex, and additional vascular risk factors and potential confounders.**

**Supplemental Table 13. All results for association of cognitive decline per 1-SD higher baseline metabolite levels with adjustment for age, sex, additional vascular risk factors and potential confounders, and stroke medications.**

**Supplemental Table 14. All results for association of cognitive decline per 1-SD higher baseline metabolite levels with adjustment for age, sex, and duration between stroke event and date blood sample was taken.**

**Supplemental Table 15. Sample selection process for genome-wide association analyses of metabolic biomarkers.**

**Supplemental Table 16. All significant results from genome-wide association analyses of metabolic biomarkers.**

**Supplemental Table 17. All Mendelian randomization results for association of metabolite levels with lacunar stroke and neuroimaging markers of small vessel disease.**

**Supplemental Table 18. Reverse Mendelian randomization results for association of lacunar stroke and neuroimaging markers of small vessel disease with metabolite levels.**

**Supplemental Table 19. Results for association of lacunar stroke and cognitive decline per 1-SD higher baseline metabolite levels with adjustment for age and sex and interaction with vascular risk factors.**

### Appendix 1

#### Members of DNA Lacunar 1 and 2 Coordinating Centres and Recruiting Centres

**DNA Lacunar 1 Study Managers:** Josie Monaghan; Alan Zanich, Samantha Febrey, Eithne Smith, Jenny Lennon, St George's University of London

**DNA Lacunar 1 Participating Centres (number of enrolled patients per centre; local investigators):** Aberdeen Royal Infirmary, Aberdeen (12; Mary Macleod). Addenbrooke's Hospital, Cambridge (54; Jean-Claude Baron, Elizabeth Warburton, Diana J Day, Julie White). Airedale General Hospital, Steeton (4; Samantha Mawer). Barnsley Hospital, Barnsley (3; Mohammad Albazzaz, Pravin Torane, Keith Elliott, Kay Hawley). Bart's and the London, London (2; Patrick Gompertz). Basingstoke and North Hampshire Hospital, Basingstoke (13; Elio Giallombardo, Deborah Dellafera). Blackpool Victoria Hospital, Blackpool (11; Mark O'Donnell). Bradford Royal Infirmary, Bradford (1; Chris Patterson). Bristol Royal Infirmary, Bristol (8; Sarah Caine). Charing Cross Hospital, London (12; Pankaj Sharma). Cheltenham General and Gloucester Royal Hospitals, Cheltenham and Gloucester (10; Dipankar Dutta). Chesterfield Royal Hospital, Chesterfield (4; Sunil Punnoose, Mahmud Sajid). Countess of Chester Hospital, Chester (22; Kausik Chatterjee). Derriford Hospital, Plymouth (4; Azlisham Mohd Nor). Dorset County Hospital NHS Foundation Trust, Dorchester (6; Rob Williams). East Kent Hospitals University NHS Foundation Trust, Kent (22; Hardeep Baht, Guna Gunathilagan). Eastbourne District General Hospital, Eastbourne (4; Conrad Athulathmudali). Frenchay Hospital, Bristol (1; Neil Baldwin). Frimley Park Hospital NHS Foundation Trust, Frimley (6; Brian Clarke). Guy's and St Thomas' Hospital, London (14; Tony Rudd). Institute of Neurology, London (25; Martin Brown). James Paget University Hospital, Great Yarmouth (1; Peter Harrison). King's College Hospital, London (16; Lalit Kalra). Leeds Teaching Hospitals NHS Trust, London (125; Ahamad Hassan). Leicester General Hospital and Royal Infirmary, Leicester (9; Tom Robinson, Amit Mistri). Luton and Dunstable NHSFT University Hospital, Luton (16; Lakshmanan Sekaran, Sakthivel Sethuraman, Frances Justin). Maidstone andTunbridge Wells NHS Trust (3; Peter Maskell). Mayday University Hospital, Croydon (14; Enas Lawrence). Medway Maritime Hospital, Gillingham (5; Sam Sanmuganathan). Milton Keynes Hospital, Milton Keynes (1; Yaw Duodu). Musgrove Park Hospital, Taunton (9; Malik Hussain). Newcastle Hospitals NHS Foundation Trust, Newcastle upon Tyne (12; Gary Ford).

Ninewells Hospital, Dundee (5; Ronald MacWalter). North Devon District Hospital, Barnstaple (8; Mervyn Dent). Nottingham University Hospitals, Nottingham (17; Philip Bath, Fiona Hammonds). Perth Royal Infirmary, Perth (2; Stuart Johnston). Peterborough City Hospital, Peterborough (1; Peter Owusu-Agyei). Queen Elizabeth Hospital, Gateshead (5; Tim Cassidy, Maria Bokhari). Radcliffe Infirmary, Oxford (5; Peter Rothwell). Rochdale Infirmary, Rochdale (4; Robert Namushi). Rotherham General Hospital, Rotherham (1; James Okwera). Royal Cornwall Hospitals NHS Trust, Truro (11; Frances Harrington, Gillian Courtauld). Royal Devon and Exeter Hospital, Exeter (22; Martin James). Royal Hallamshire Hospital, Sheffield (1; Graham Venables). Royal Liverpool University Hospital and Broadgreen Hospital, Liverpool (9; Aravind Manoj). Royal Preston Hospital, Preston (18; Shuja Punekar). Royal Surrey County Hospital, Guildford (23; Adrian Blight, Kath Pasco). Royal Sussex County Hospital, Brighton (14; Chakravarthi Rajkumar, Joanna Breeds). Royal United Hospital, Bath (6; Louise Shaw, Barbara Madigan). Salford Royal Hospital, Salford (16; Jane Molloy). Southampton General Hospital, Southampton (1; Giles Durward). Southend Hospital, Westcliff-on-Sea (26; Paul Guyler). Southern General Hospital, Glasgow (34; Keith Muir, Wilma Smith). St George’s Hospital, London (108; Hugh Markus). St Helier Hospital, Carshalton (10; Val Jones). Stepping Hill Hospital, Stockport (4; Shivakumar Krishnamoorthy). Sunderland Royal Hospital, Sunderland (1; Nikhil Majumdar). The Royal Bournemouth Hospital, Bournemouth (15; Damian Jenkinson). The Walton Centre, Liverpool (15; Richard White). Torbay Hospital, Torquay (19; Debs Kelly). University Hospital Aintree, Liverpool (19; Ramesh Durairaj). University Hospital of North Staffordshire, Stoke-on-trent (16; David Wilcock). Wansbeck General Hospital and North Tyneside Hospital, Ashington and North Shields (6; Christopher Price). West Cumberland Hospital, Whitehaven (6; Olu Orugun, Rachel Glover). West Hertfordshire Hospital, Watford (20; David Collas). Western General Hospital, Edinburgh (12; Cathie Sudlow). Western Infirmary, Glasgow (33; Kennedy R. Lees, Jesse Dawson). Wycombe Hospital and Stoke Mandeville, High Wycombe (20; Dennis Briley and Matthew Burn). Yeovil District Hospital, Yeovil (46; Khalid Rashed). York Teaching Hospital, York (1; John Coyle).

**DNA Lacunar 2 Coordinating Centre:** Chief Investigator: Hugh Markus; Trial Coordinators: Rebecca Dowden, Laurence Loubiere, Elnara Aghakishiyeva; Case Reviewer: Lupei Cai.

**DNA Lacunar 2 Recruiting Centres (local investigator, number of recruited participants as of 31 January 2025):** Addenbrookes Hospital (Hugh Markus, 299), James Paget Hospital (Carlo Canepa, 3), Peterborough City Hospital (Santhosh Subramonian, 13), North Middlesex

Hospital (Robert Luder, 12), Royal Stoke Hospital (Janaka Weerathunga, 88), Royal London Hospital (Farhad Huwez, 21), West Suffolk Hospital (Abul Azim, 89), Watford General Hospital (Mohit Bhandari, 99), Royal Cornwall Hospital (Katja Adie, 20), New Cross Hospital, Wolverhampton (Nasar Ahmad, 34), Princess Royal University Hospital, Telford (Uttam Sinha, 2), King's Mill Hospital (Jahanzeb Rehan, 27), King's College Hospital (James Teo, 42), Queen Elizabeth Hospital, King's Lynn (Umesh Rai, 4), Countess of Chester Hospital (Kausik Chatterjee, 23), Leeds General Infirmary (Ahamad Hassan, 53), Royal Hallamshire Hospital (Aaizza Naqvi, 18), Wirral University Teaching Hospital (Ruth Davies, 0), University Hospital of North Tees (Ijaz Anwar, 12), Lister Hospital (Aparna Pusalkar, 4), Royal Bournemouth Hospital (Kamy Thavansan, 32), Royal Hampshire County Hospital (Nigel Smyth, 14), Derriford Hospital, Plymouth (Azlisham Mohd Nor, 25), Torbay Hospital (Biju Bhaskaran, 2), Pinderfields Hospital (Prabal Datta, 10), Queen's Hospital, Havering (Abhijit Chaudhuri, 0), Royal Devon and Exeter Hospital (Martin James, 15), University College Hospital (David Werring, 84), Nottingham City Hospital (Kailash Krishnan, 28), Salford Royal Hospital (Dwaipayan Sen, 11), James Cook University Hospital, South Tees (Samer Al-Hussayni, 31), University Hospital of North Durham (Revin Thomas, 52), Yeovil District Hospital (Khalid Rashed, 63), St Peter's Hospital (Brendan Affley, 29), Sandwell General Hospital (David Gull, 21), Guys and St Thomas' Hospital (Ajay Bhalla, 48), Royal Sussex County Hospital (Chakravarthi Rajkumar, 1), Southampton General Hospital (Emma Battersby-Wood, 31), Doncaster Royal Infirmary (Dinesh Chada, 26), St George's Hospital (Usman Khan, 123), Wycombe Hospital (Matthew Burn, 20), Musgrove Park Hospital (Malik Hussain, 37), Scunthorpe General Hospital (Amit Banerjee, 3), Ipswich Hospital (Sajid Alam, 17), Southend University Hospital (Paul Guyler, 12), Gloucestershire Royal Hospital (Dipankar Dutta, 23), Queen Elizabeth The Queen Mother Hospital, Margate (Saidu Abubakar, 9), Colchester General Hospital (Rajesh Saksena, 34), Royal Derby Hospital (Tim England, 5), Croydon University Hospital (Enas Lawrence, 66), Norfolk and Norwich University Hospital (Kneale Metcalf, 12), Leicester Royal Infirmary (Amit Mistri, 16), William Harvey Hospital (Ibrahim Balogun, 5), Medway Maritime Hospital (Samuel Sanmuganathan, 5), Kent and Canterbury Hospital (Saidu Abubakar, 16), Sunderland Royal Hospital (Naweed Sattar, 18), University Hospital Llandough (Benjamin Jelley, 11), Cheltenham General Hospital (Dipankar Dutta, 0), Royal Victoria Infirmary (Akif Gani, 16), University Hospital Coventry & Warwickshire (Munib Mirza, 16), Fairfield General Hospital (Narayanamoorthi Saravanan, 14), Milton Keynes University Hospital (Janet Costa, 9), Royal Blackburn Hospital (Anand Nair, 1), Royal Preston Hospital (Hedley Emsley, 13), Stepping Hill Hospital (Appukuttan

Suman, 6), Royal Albert Edward Infirmary (Habib Rehman, 12), Broomfield Hospital (Ramanathan Kirthivasan, 34), Basildon University Hospital (Jigisha Amin, 1), Homerton University Hospital (Thomas Harrison, 0), Queen Elizabeth Hospital, Gateshead (Louise Southern, 0), and Southmead Hospital (Sandeep Buddha, 0).

### Supplemental Methods

**Detailed Methods. Further details of genome-wide association analyses.**

We conducted a genome-wide association analysis for each of the 250 metabolites using genetic data from DL1 and DL2. Genotyping was conducted by University College London Genomics at the Zayed Centre for Research into Rare Disease in Children using the Infinium Global Screening Array-24 kit. Samples with a low genotype call rate (< 0.90) were removed, and the genotyped datasets were harmonized against the reference panel 'PASS.Variantsbravo-dbsnp-all_hg37.tab.gz' and pre-imputation quality control (QC) filters were applied. Variants were excluded if they were missing from a substantial number of samples (≥ 3%), if they had low minor allele frequencies (< 1%) or Hardy-Weinberg equilibrium *P*-values ($< 1*10^{-7}$), or if they were non-autosomal. Samples were also excluded if they had a substantial number of missing SNPs (≥ 3%). Imputation was performed on the TOPMed Imputation Server against the TOPMed R2 reference panel utilizing Minimac4.[25] Imputed SNPs with sufficiently high imputation scores (Rsq > 0.3) were kept. Imputed genotypes were sorted and stored according to their chromosomal positions on build GRCh38.

Post-imputation harmonization was carried out using Genotype Harmonizer[26] and post-imputation QC filters were applied. Samples were excluded if they were genotyped for less than 95% of SNPs, and variants were excluded if they were genotyped in less than 97% of samples or if their allele frequencies were less than 0.5%. Kinship analysis was performed with the KING algorithm in PLINK v2 and pi-hat test in PLINK v1.9,[27] and one sample from each pair of samples with a kinship coefficient > 0.345 or pi-hat test > 0.2 was removed from further analysis. LD pruning was carried out to remove highly correlated SNPs and a principal component analysis (PCA) was performed on the pruned dataset to identify the top 20 principal components for genetic ancestry.

Association testing was conducted using Regenie v3.4.1[28] with adjustment for age, sex, and the top three principal components for ancestry, and the GWAS results were visualized using Manhattan and QQ plots. A false discovery rate (FDR) threshold of 5% was applied, and the FDR-corrected significantly associated variants were selected. Linkage disequilibrium (LD) clumping using a threshold of $r^2 < 0.5$ was applied, and the independent SNPs for each metabolite were selected as genetic instruments for the Mendelian randomization analyses.

### Supplemental Figures

**Supplemental Figure 1. Volcano plot showing the results of cross-sectional univariate analyses for the association between 250 metabolic biomarkers and lacunar stroke.**

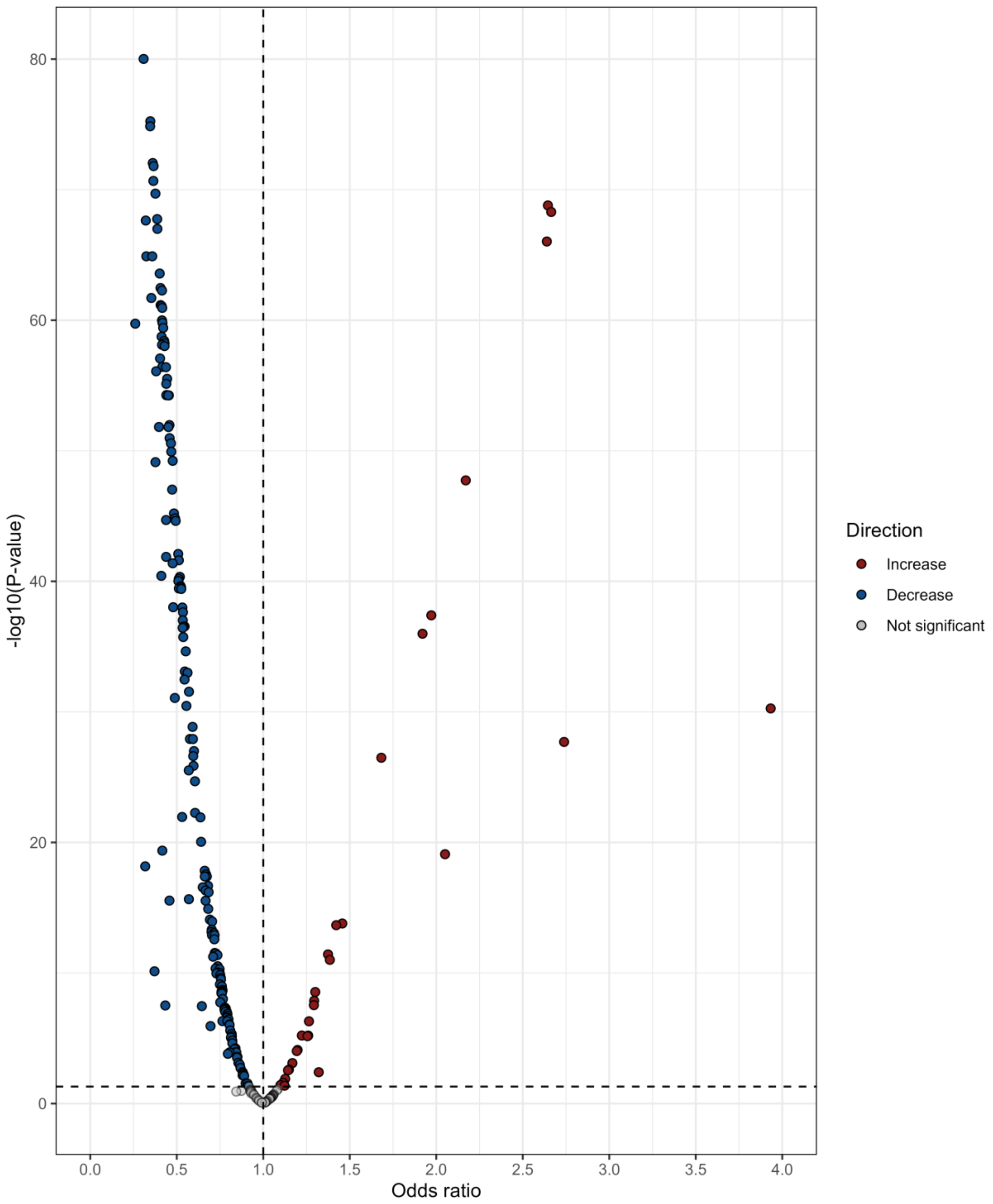


Odds ratios are shown on the x-axis and -$\log_{10}$ *P*-values are shown on the y-axis. The FDR-corrected threshold of $P < 0.05$ is shown as a dashed horizontal line. Metabolites significantly associated with increased risk of lacunar stroke are shown in red and metabolites significantly associated with decreased risk of lacunar stroke are shown in blue.

**Supplemental Figure 2. Circular bar plot showing the results of cross-sectional univariate analyses for the association of 250 metabolic biomarkers with ILI cases versus controls.**

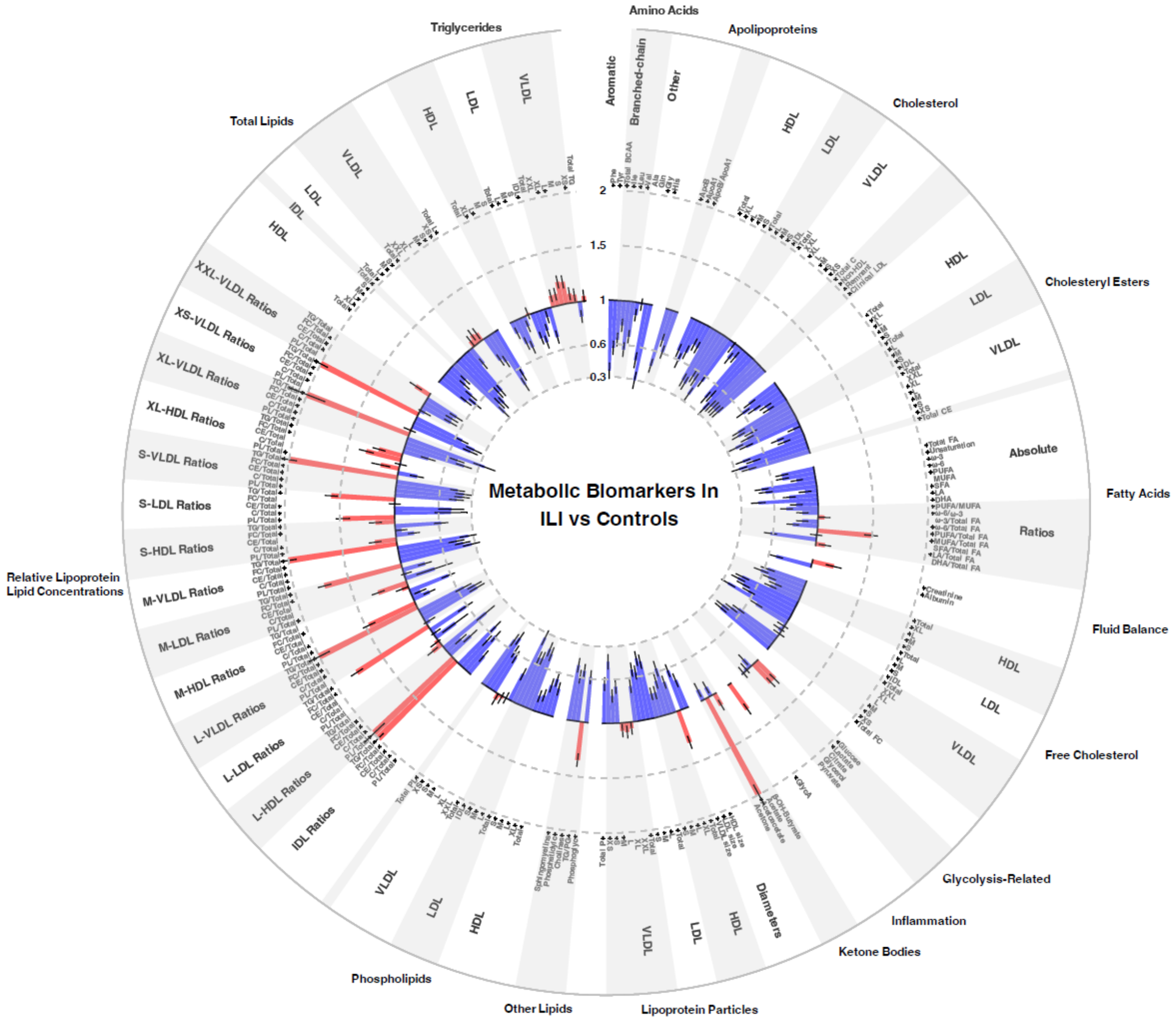


Abbreviations: FDR: false discovery rate; ILI: isolated lacunar infarcts; SD: standard deviation. Odds ratios with 95% confidence intervals are presented for the association with ILI versus controls per 1-SD increase in metabolite concentration with adjustment for age, sex, hypertension, and recruitment centre. Odds ratios >1 showing positive associations are represented in red, and odds ratios <1 showing inverse associations are in blue. Associations significant at an FDR-corrected $P < 0.05$ are indicated with an asterisk (*) alongside the names of the respective metabolites.

**Supplemental Figure 3. Circular bar plot showing the results of cross-sectional univariate analyses for the association of 250 metabolic biomarkers with MLI/LA cases versus controls.**

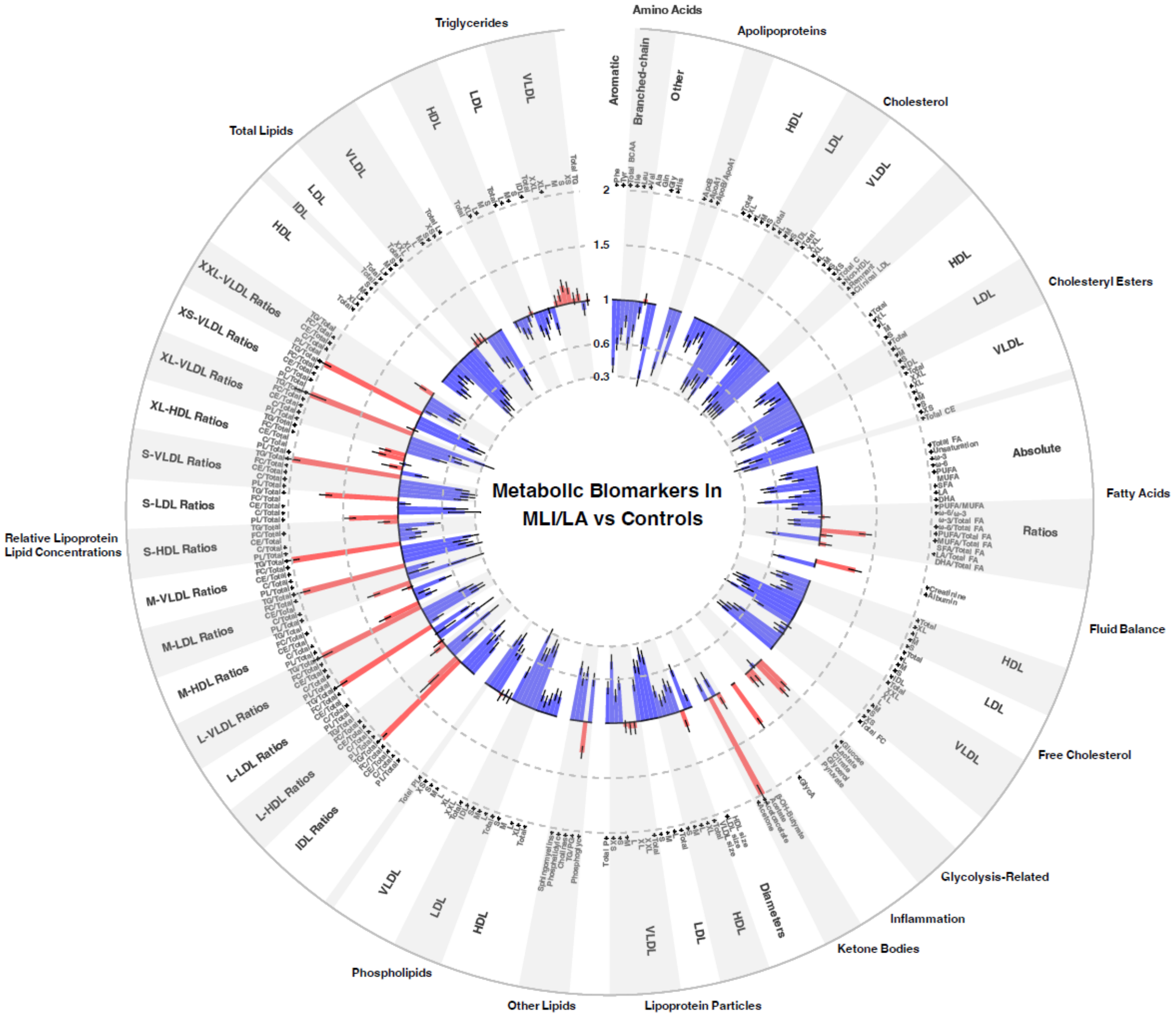


Abbreviations: FDR: false discovery rate; MLI: multiple lacunar infarcts; LA: leukoaraiosis; SD: standard deviation. Odds ratios with 95% confidence intervals are presented for the association with MLI or LA versus controls per 1-SD increase in metabolite concentration with adjustment for age, sex, hypertension, and recruitment centre. Odds ratios >1 showing positive associations are represented in red, and odds ratios <1 showing inverse associations are in blue. Associations significant at an FDR-corrected $P < 0.05$ are indicated with an asterisk (*) alongside the names of the respective metabolites.

**Supplemental Figure 4. Scree plot and loadings plot from principal component analysis of metabolite levels. (A) Scree plot. (B) Loadings plot.**

**(A) Scree plot.**

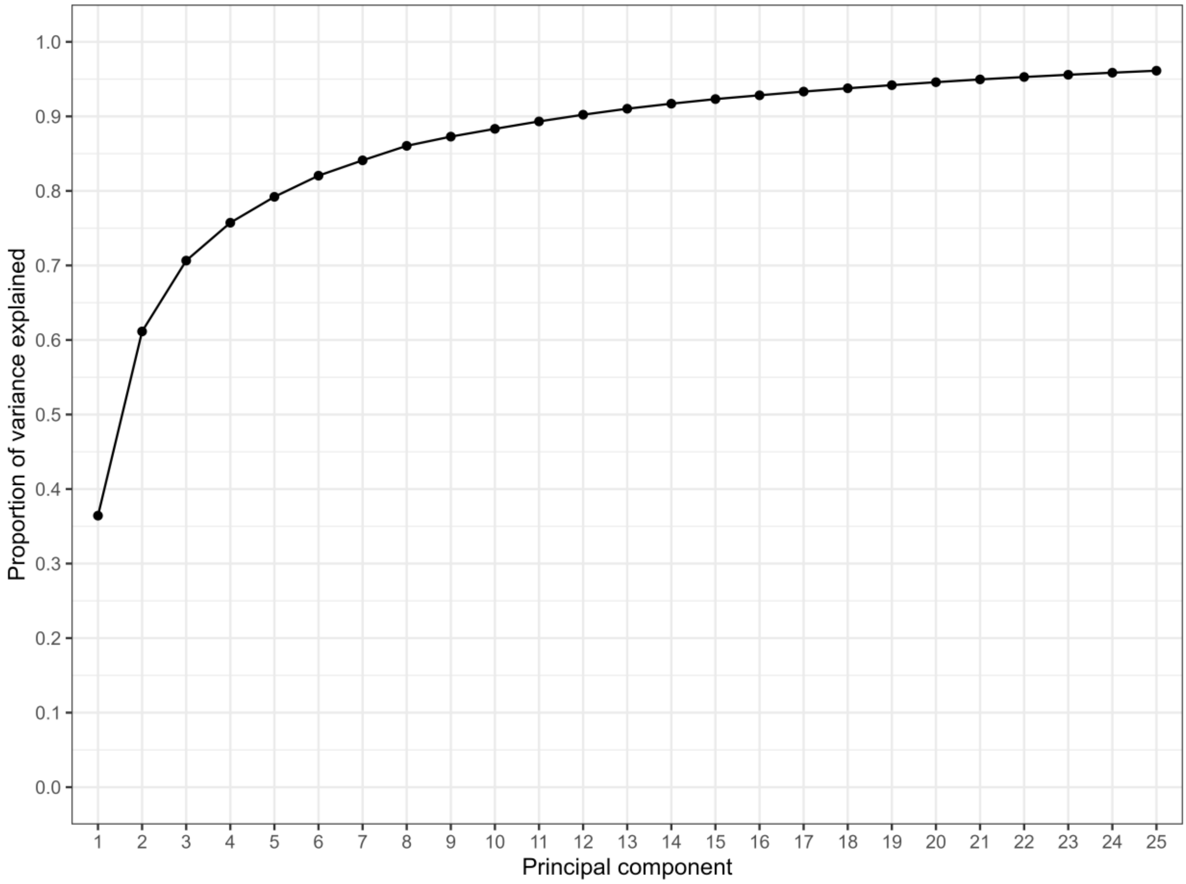

**(B) Loadings plot.**

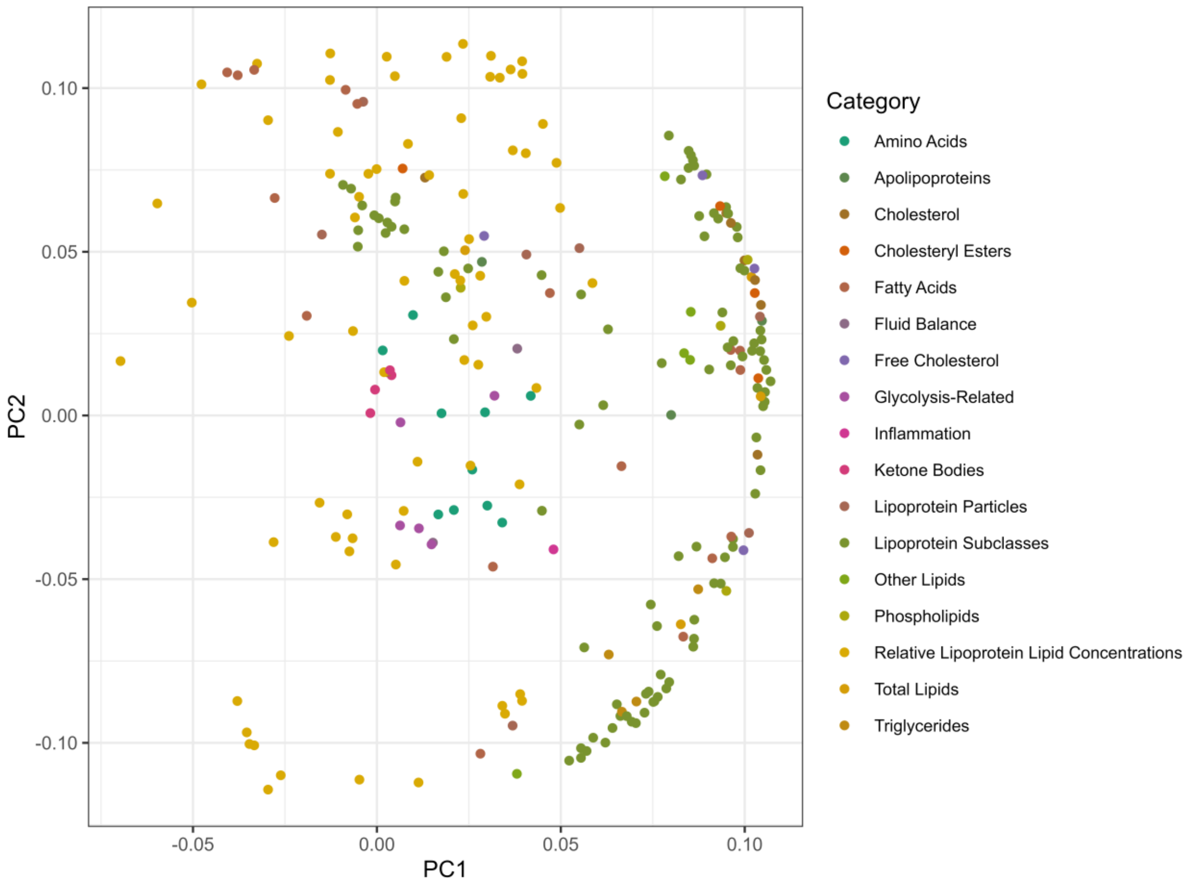


The scree plot **(A)** shows the cumulative proportion of variance explained by each successive principal component of the metabolite concentrations. The loadings plot **(B)** shows a scatter plot of the matrix loadings for the first and second principal components of the metabolite concentrations.

**Supplemental Figure 5. Forest plot showing the results of cross-sectional univariate analyses for the association of the first twenty-two principal components of metabolite levels with lacunar stroke status, ILI vs controls, and MLI/LA vs controls.**

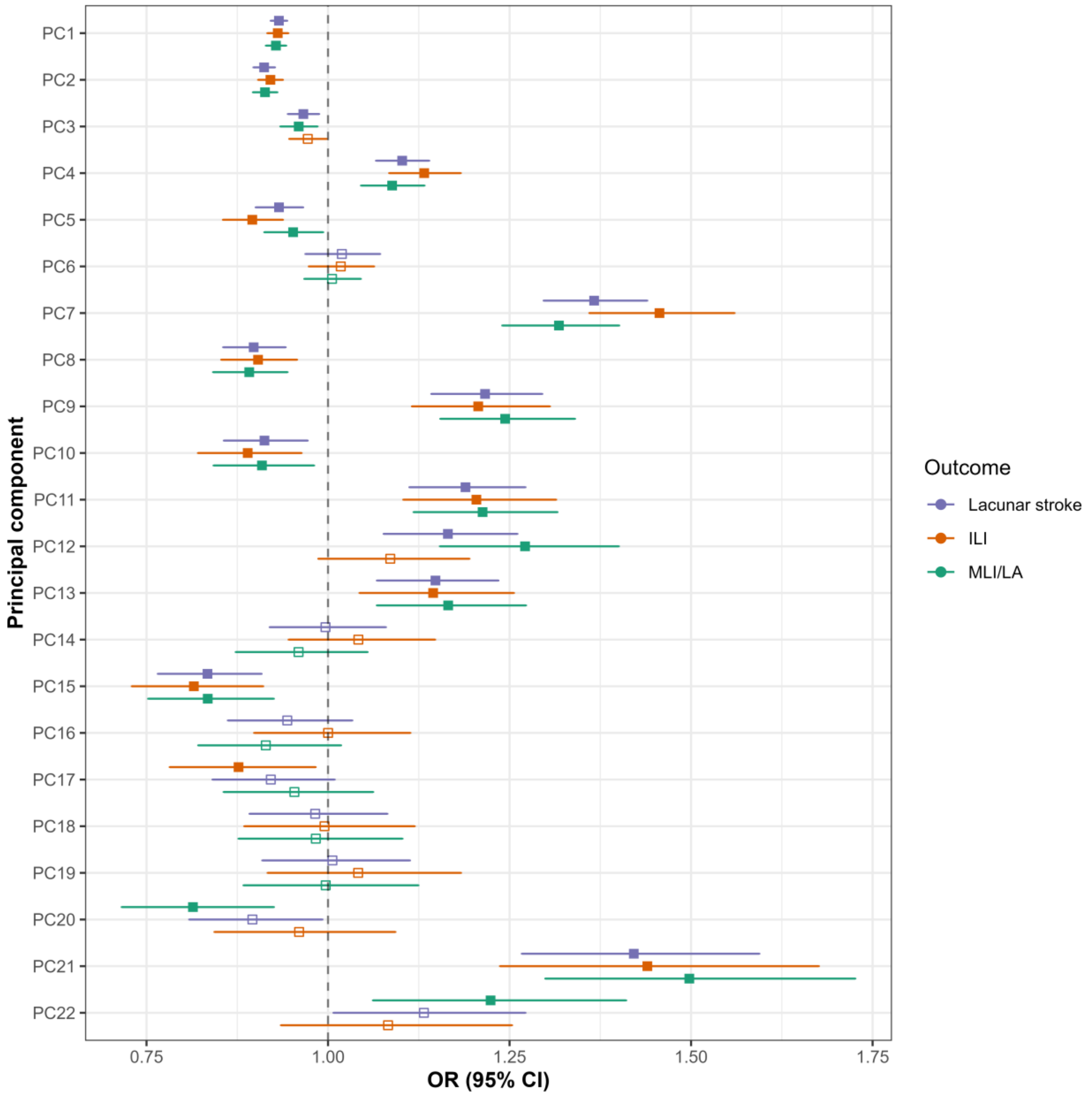


Abbreviations: CI: confidence interval; FDR: false discovery rate; ILI: isolated lacunar infarcts; MLI: multiple lacunar infarcts; LA: leukoaraiosis; OR: odds ratio. Odds ratios with 95% confidence intervals for the association with lacunar stroke status, ILI vs controls, and MLI/LA vs controls are presented per 1-SD increase in metabolite concentration with adjustment for age and sex. Associations significant at an FDR-corrected $P < 0.05$ are indicated with filled points while associations that are not statistically significant are shown with hollow points.

**Supplemental Figure 6. Scatter plots of genetic associations of metabolic levels with lacunar stroke and neuroimaging markers of SVD for Mendelian randomization association analyses with significant or suggestive associations.**

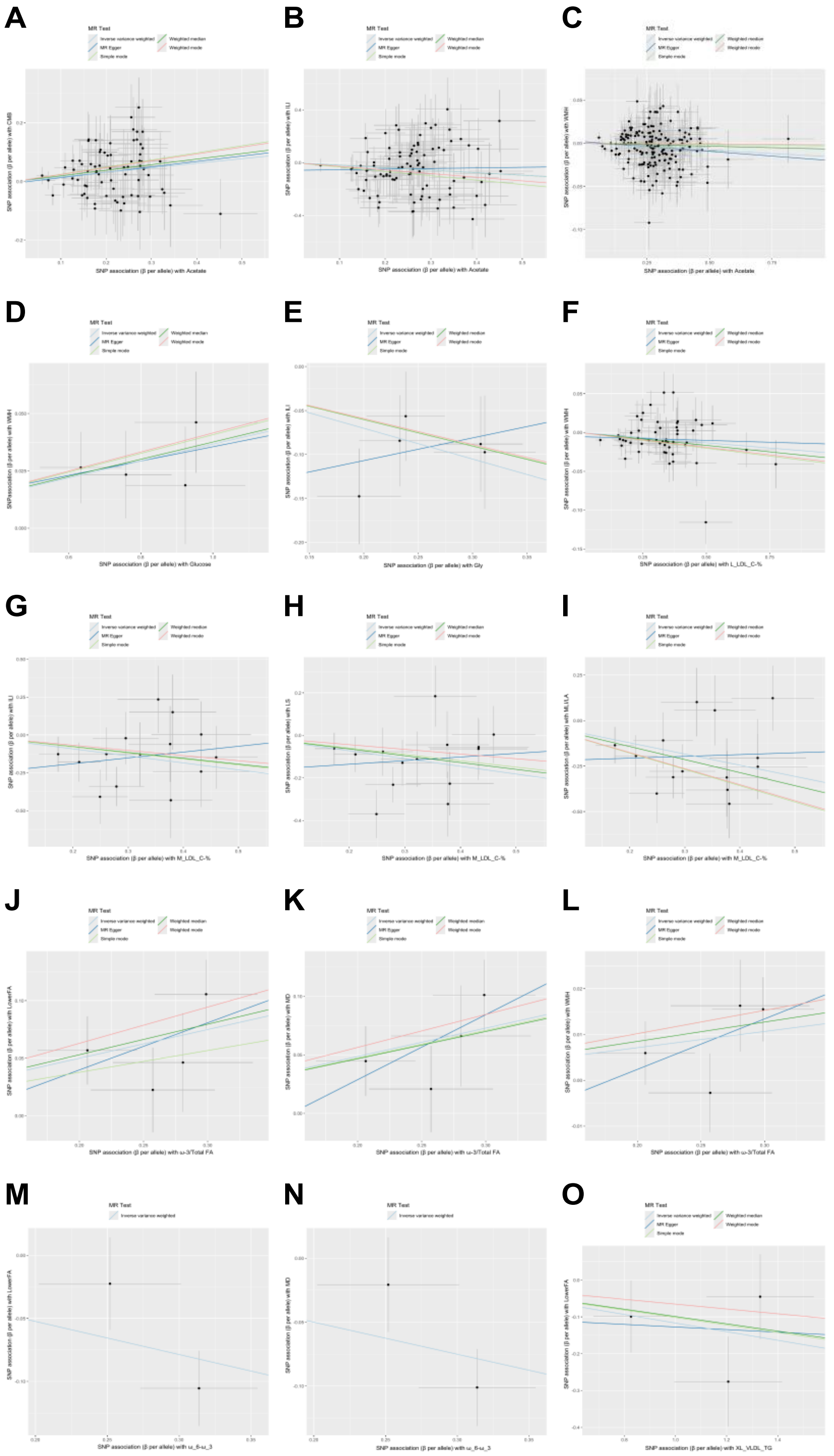

Abbreviations: MR: Mendelian randomization; SNP: single nucleotide polymorphism; SVD: small vessel disease. Scatter plots are shown for the Mendelian randomization results with evidence of a significant or suggestive association (at an uncorrected $P < 0.05$) between metabolites and lacunar stroke and MRI markers of SVD using the inverse-variance weighted or Wald ratio method. The x-axis of each plot shows the association of each genetic variant with the metabolite, and the y-axis shows the association of each genetic variant with the outcome. The slope of each line corresponds to an estimate of the causal effect using the various MR methods.
